# Cryo-PRO facilitates whole blood cryopreservation for single-cell RNA sequencing of immune cells from clinical samples

**DOI:** 10.1101/2024.09.18.24313760

**Authors:** Alyssa K. DuBois, Pierre O. Ankomah, Alexis C. Campbell, Renee Hua, Olivia K. Nelson, Christopher A. Zeuthen, M. Kartik Das, Shira Mann, Abigail Mauermann, Blair A. Parry, Nathan I. Shapiro, Michael R. Filbin, Roby P. Bhattacharyya

## Abstract

Single-cell RNA sequencing (scRNA-seq) of peripheral blood mononuclear cells (PBMCs) has enhanced our understanding of host immune mechanisms in small cohorts, particularly in diseases with complex and heterogeneous immune responses such as sepsis. However, standard PBMC isolation from blood requires technical expertise and over two hours of onsite processing using Ficoll density gradient separation (‘Ficoll’) for scRNA-seq compatibility, precluding large-scale sample collection at most clinical sites. To minimize onsite processing, we developed Cryo-PRO (Cryopreservation with PBMC Recovery Offsite), a method of immediate onsite whole blood cryopreservation and subsequent batched PBMC isolation in a central laboratory prior to sequencing. We compared multimodal single-cell immune profiling results from samples processed using Cryo-PRO versus standard onsite Ficoll separation in 23 patients with sepsis. Critical outputs including cell substate fractions, marker genes, and surface protein expression were similar for each method across multiple cell types and substates, including an important monocyte substate enriched in patients with sepsis. Capture of T cell receptor transcripts was also comparable across both methods. Cryo-PRO reduced onsite sample processing time from >2 hours to <15 minutes and was reproducible across two enrollment sites, thus demonstrating potential for expanding multimodal single-cell analyses in multicenter studies of sepsis and other diseases.

## INTRODUCTION

Single-cell RNA sequencing (scRNA-seq) is a pivotal methodology with great potential for advancing the understanding of biological systems (1). It has revealed previously unrecognized cell types and transcriptional substates within complex tissues and demonstrated differences in gene expression that illuminate pathophysiological variation in many aspects of human disease (2–4). ScRNA-seq has particular appeal for dissecting the cellular basis for heterogeneity in diseases and opening pathways to precision medicine. Sepsis, for example, is a syndrome with expansive differences in clinical course and outcome between patients that is impacted substantially by heterogeneity in patients’ immunological responses. However, there has been limited progress in understanding this variation using existing methods. In particular, transcriptional profiling in patients with sepsis has mostly relied on bulk RNA sequencing (bulk RNA-seq) to generate averaged signatures from all circulating immune cells, obscuring the cellular basis and underlying mechanisms of immune dysfunction (5–9).

In previous work, our group has used scRNA-seq to profile circulating peripheral blood mononuclear cells (PBMCs) in urosepsis, identifying a unique CD14+ monocyte subtype (monocyte substate 1, or MS1), that is expanded in sepsis relative to infection without sepsis (10). These monocytes have a gene expression profile similar to myeloid-derived immune suppressor cells, which are immune regulatory cells that inhibit T cell activation, proliferation and cytotoxic activity (11). MS1 cells may therefore play an immunosuppressive role in sepsis and contribute to an important transcriptional subphenotype, or “endotype”, of sepsis (12). Several other studies have employed scRNA-seq in small sepsis cohorts (12–15). However, resolution of sepsis endotypes and the contributory roles of immune cell subtypes requires large cohorts enrolled at multiple, geographically-separated clinical sites. Such large-scale studies are necessary to appropriately represent the heterogeneity of sepsis with its variable pathogen types, anatomic sites of infection, timing of presentation, severity, trajectory, clinical characteristics (e.g., age, sex, race and ethnicity, comorbidities), and complex host immune responses. Leveraging the resolution of scRNA-seq at a scale that allows sufficient sampling of all relevant subphenotypes of sepsis has the potential to enable endotyping assessment with sufficient accuracy to impact sepsis care.

Implementing scRNA-seq studies in clinical settings at scale is challenged by several logistical barriers. Blood, which offers a diverse and dynamic snapshot of the systemic response to infection, serves as a key resource for investigating immune responses in sepsis and other conditions (16). However, because live blood cells are highly sensitive to environmental perturbations, it is necessary to either process samples rapidly before sequencing or employ cryostorage for later analysis to minimize *ex vivo* transcriptional changes after blood collection. Therefore, processing the blood sample to a point where transcription is halted (e.g., by freezing live cells or fixing them unless sequencing is performed immediately) often falls to operators at the collection site. Currently, standard practice for scRNA-seq studies of PBMCs involves a density gradient centrifugation step immediately following blood draw (Ficoll-paque processing, or “Ficoll”) to isolate and store immune cells (17). This process, which must be completed promptly at the site of blood collection, is resource-intensive, time-consuming, sensitive to protocol variations, and requires technical skill and training. The complexity of real-time processing of whole blood samples has limited the widespread use of scRNA-seq in clinical investigations. Moreover, the lack of standardization in processing and analysis can lead to batch effects, hindering comparisons across sites and between studies.

To overcome the practical limitations of scRNA-seq in clinical settings, we developed Cryo-PRO (Cryopreservation with PBMC Recovery Offsite), a method for isolating PBMCs from cryopreserved whole blood samples. The approach utilizes magnetic depletion of red blood cells followed by fluorescence-activated cell sorting to recover immune cells for scRNA-seq. Cryo-PRO enables the immediate cryopreservation of whole blood samples at clinical sites, with onsite freezing and storage, allowing for their transfer at a later time to a centralized laboratory for PBMC isolation and single-cell profiling. In this study, we directly compare the single-cell transcriptome (scRNA-seq), surface proteome, and T cell receptor sequencing (TCR-seq) output from sepsis patient samples processed using Cryo-PRO with those processed by the standard onsite Ficoll-gradient separation method. Our findings demonstrate technical equivalence and biological reproducibility between the two methods. Cryo-PRO has the potential to enable broad application of multimodal single-cell analysis to multicenter studies and clinical trials by simplifying sample collection and centralizing cell isolation to improve cost efficiency, minimize batch effects, and increase sample sizes. This method could advance our understanding of the complexity of sepsis and other heterogeneous diseases, enabling development of precision diagnostics and targeted therapeutic strategies.

## RESULTS

Patients greater than 18 years of age who presented to the Emergency Departments of two large, academic hospitals in Boston, Massachusetts — Massachusetts General Hospital (MGH) and Beth Israel Deaconess Medical Center (BIDMC) — with clinical concern for sepsis or septic shock with associated organ dysfunction were enrolled in the study. Up to 10 mL of blood was obtained from patients and processing was initiated onsite using two methods: 1) standard Ficoll gradient separation from whole blood by following standard procedures for isolating and freezing PBMCs (17), followed by storage at -80°C; and 2) Cryo-PRO, by immediately adding dimethyl sulfoxide (DMSO) to a final concentration of 10% (v/v) in 1 mL aliquots of fresh whole blood and freezing at -80°C. To measure potential effects of variation in processing by enrollment site, for a subset of patients, up to 20 mL of blood (separated in two 10-mL tubes) was obtained; one tube was immediately couriered to the other clinical site while one tube remained at the enrolling site. Processing at both sites using both Ficoll and Cryo-PRO began at the same time upon sample receipt at the receiving site. Blood from one healthy donor was obtained from Research Blood Components (Watertown, MA) and processed using both Ficoll and Cryo-PRO methods to use as a reference standard. All samples were sent to the Eli and Edythe L. Broad Institute of MIT and Harvard (Broad Institute) for long term storage at -140°C, subsequent processing, and multimodal single-cell immune profiling.

We adjudicated 23 subjects based on the presence of sepsis or bacterial infection for sample processing and sequencing. Septic shock requiring vasopressors for over 24 hours was present in 15 subjects (65%), sepsis without shock in 6 subjects (26%), and bacterial infection not meeting Sepsis-3 criteria (18) in 2 subjects (9%). Bacteremia was present in 7 (30%) of the 23 subjects. The median patient age was 66 years (IQR 62.5 - 76.5), with 35% female. The healthy donor was a 63 year old male.

Of the 23 included subjects, 8 were processed in parallel at both sites and 15 were processed only at the site of enrollment. Patient-paired frozen Ficoll and Cryo-PRO samples were processed for single-cell analysis at the Broad Institute. Processing included a magnetic red blood cell depletion step (Cryo-PRO samples only), fluorescence-activated cell sorting to recover live PBMCs (DAPI-CD45+ CD235a-CD15-cells), and a standard workflow for droplet-based single-cell RNA capture with surface proteome and T cell receptor repertoire measurements (10X Genomics Chromium Next GEM 5’ v2 Kit with cellular indexing of transcriptional epitope sequencing (CITE-seq)) (see Methods) (19). Sample hashing was used to enable pooling of eight samples per processing batch, and to facilitate post-sequencing demultiplexing and multiplet detection. An overview of the sample collection, storage, and processing strategies is summarized in Figure 1.

**Figure 1.**
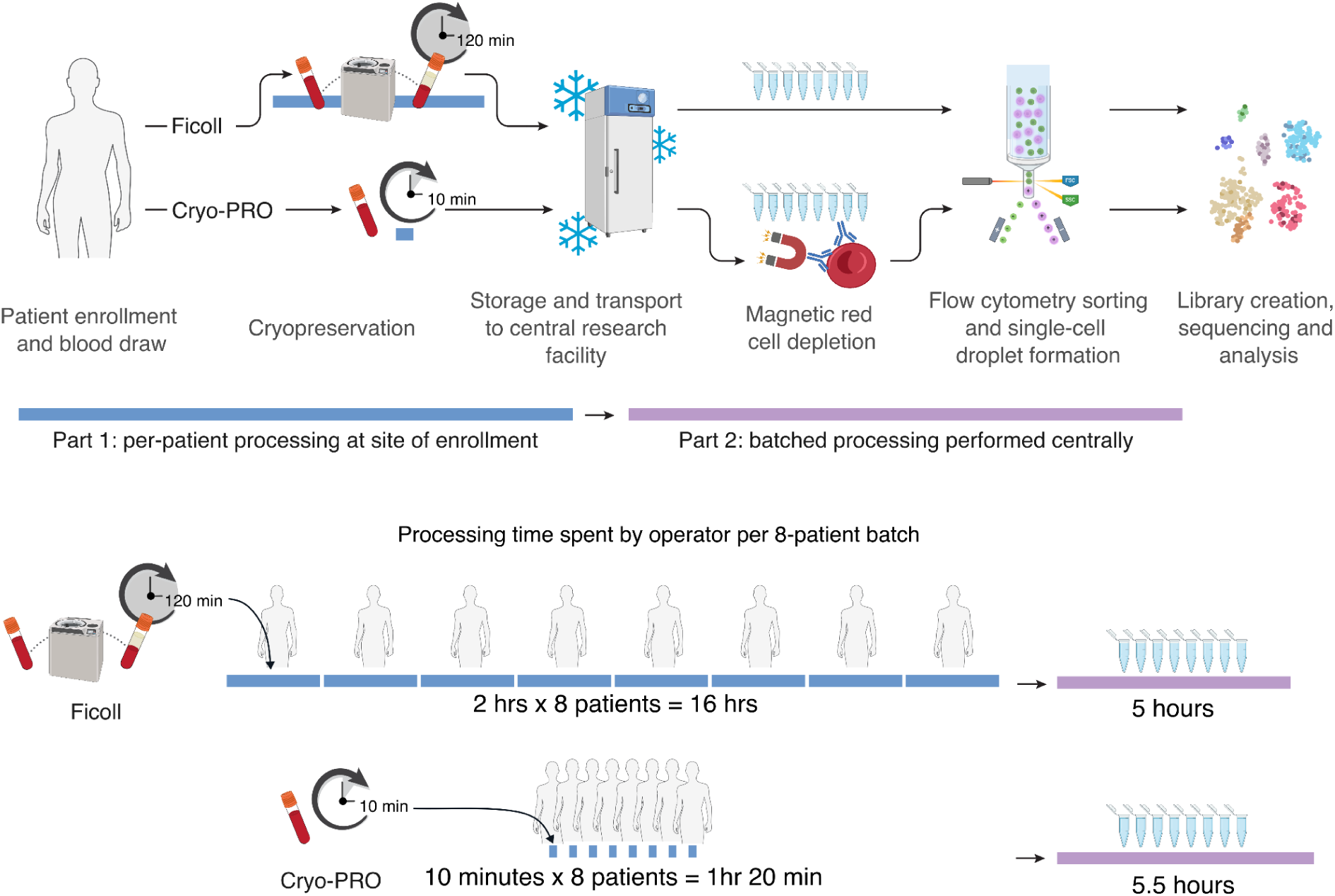
Overview of Ficoll and Cryo-PRO sample processing workflows. Cryo-PRO is designed to expedite sample processing at the site of collection by incorporating a whole blood cryopreservation step (and subsequent red cell depletion step) to replace standard Ficoll processing. Blue bars denote processing done at site of enrollment; purple bars denote processing done at a centralized laboratory and include steps up to single-cell droplet formation.

### Samples processed using Cryo-PRO yield high quality scRNA-seq and surface proteome data with minimal on-site processing time

The mean time required for complete on-site processing (from processing start time to storage at -80°C) for Ficoll samples was 2 hours and 23 minutes (SD: 40 minutes), while Cryo-PRO samples required an average of 13 minutes (SD: 7 minutes) (Figure 2A). The proportion of viable PBMCs recovered by either method was estimated using live/dead staining during flow cytometry sorting. The mean proportion of live (DAPI-) PBMCs (CD235a-CD15-CD45+ cells) was 96.7% (SD 3.0%) for Ficoll samples and 94.1% (SD 8.4%) for Cryo-PRO samples (Figure 2B).

**Figure 2.**
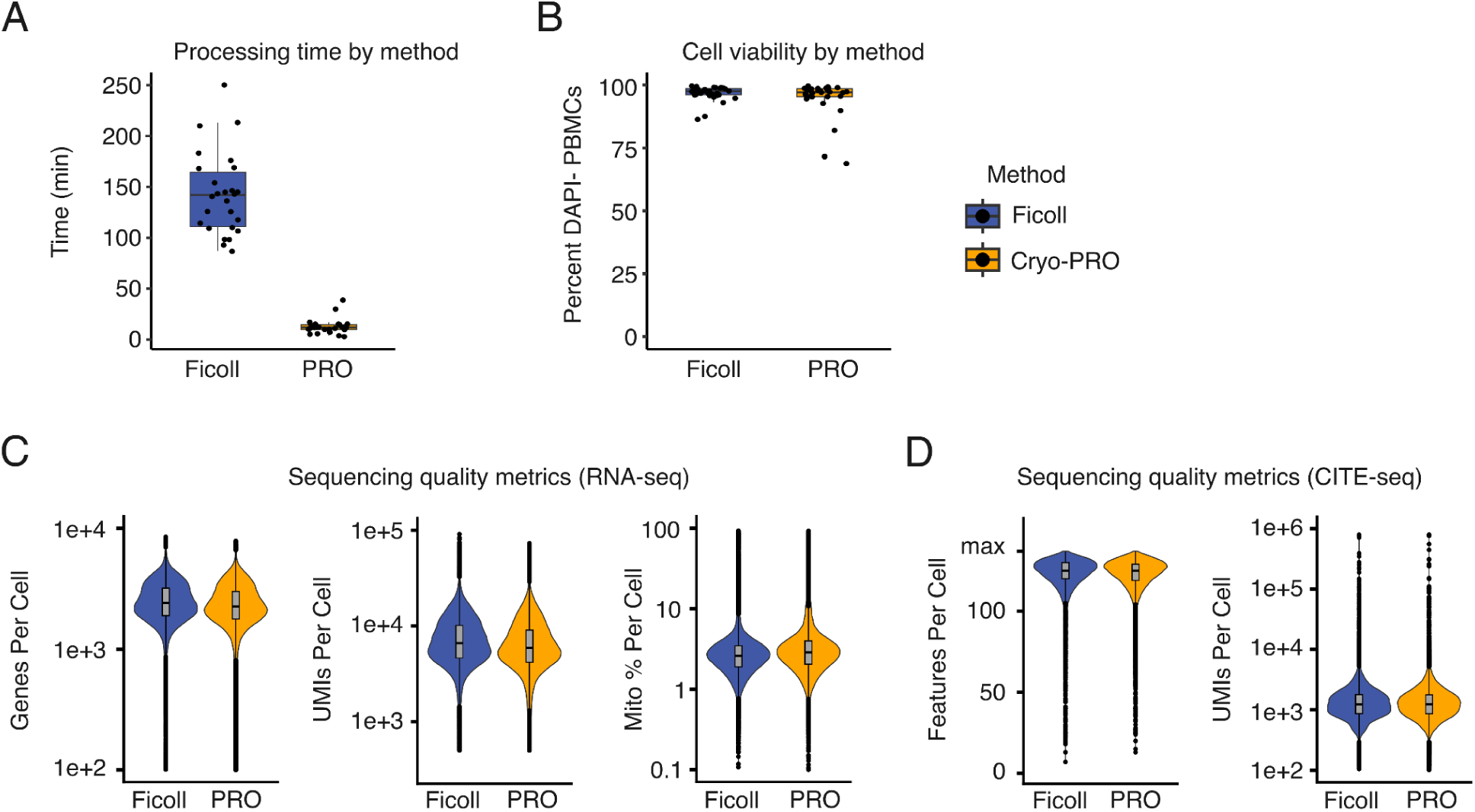
Processing time and quality metrics by cryopreservation method. (A) “Hands-on” time spent by operators at clinical sites to process patient samples from initiation of processing after blood draw to placing the sample in a freezer for storage. (B) Percent of CD45+ CD235a-CD15-cells staining DAPI negative on flow cytometry as an indicator of cell membrane integrity and cell viability. Boxplots show the median (center line) and interquartile range (box); whiskers extend to the most extreme values within 1.5-fold the interquartile range (IQR). Points represent individual samples. (C) Violin plots of RNA sequencing quality metrics by method (left to right): unique genes per cell, unique molecular identifiers (UMIs) of RNA transcripts per cell, percent of transcripts represented by mitochondrial genes per cell. (D) Violin plots of CITE-seq quality metrics by method: unique surface protein features (left panel) and UMIs (right panel) per cell (detected via CITE-seq). Violin plots show the distribution of each metric detected per cell; embedded boxplots indicate the median and IQR (whiskers = 1.5x IQR). PRO denotes Cryo-PRO.

The Cellranger pipeline (10X Genomics) was used to process the raw sequencing data, and the Seurat V5 package in R (20) was used for subsequent analysis of single-cell sequencing data (see Methods). Multiplets (cells associated with more than one patient hashtag) were removed from analysis. We recovered an average of 2,690 (SD 950) and 2,472 (SD 918) singlet cells per sample for the Ficoll and Cryo-PRO methods respectively (Supplemental Figure 1). Sequencing quality was assessed using the following standard metrics: 1) number of genes sequenced per cell, 2) number of unique molecular identifiers (UMIs) per cell, and 3) percent of mitochondrial genes sequenced per cell. Higher numbers of genes per cell and UMIs per cell indicate greater per-cell transcript recovery, while a greater percentage of mitochondrial genes suggests cell damage (21). Quality metrics showed similar distributions between methods (Figure 2C). The majority of cells (97.6% from Ficoll processing and 94.4% from Cryo-PRO) passed commonly-used quality thresholds (>250 genes per cell, >1,000 UMIs, and <10% mitochondrial genes) (22). Detection of antibody-derived tags (ADT) used for CITE-seq surface proteome measurement was similar between the two methods (Figure 2D). Quality metrics were similar between methods at an individual patient level (Supplemental Figure 2).

### Cryo-PRO enables identification of transcriptional substates, gene expression patterns, and surface protein markers

We next assessed whether Cryo-PRO supports scRNA-seq and surface proteome datasets of sufficient quality to reproduce biologically relevant results compared to Ficoll. ScRNA-seq analysis was performed separately for cells obtained from each processing method (86,083 cells for Ficoll and 79,089 cells for Cryo-PRO) to ensure independent identification of cell identity and gene expression patterns (see Methods). Clusters enriched for dead and dying cells, identified by the predominance of mitochondrial genes, were removed from further analysis as an additional quality control measure (23). We identified transcriptionally similar cells that expressed canonical marker genes for the major mononuclear immune cell types (i.e., T cells, B cells, natural killer cells, monocytes, and dendritic cells). Subclustering within each cell type identified higher-resolution clusters of cells with additional transcriptional similarity (i.e., cell substates, e.g., CD4+ memory T cells, naive B cells, etc.), which were classified by comparison with reference datasets (20). All the major mononuclear immune cell types, comprising a total of 17 cell substates, were identified from cells isolated using either Ficoll or Cryo-PRO, each clustered independently but projected onto a shared set of two-dimensional uniform manifold approximation and projection (UMAP) axes for visualization (Figure 3A; see Methods).

**Figure 3.**
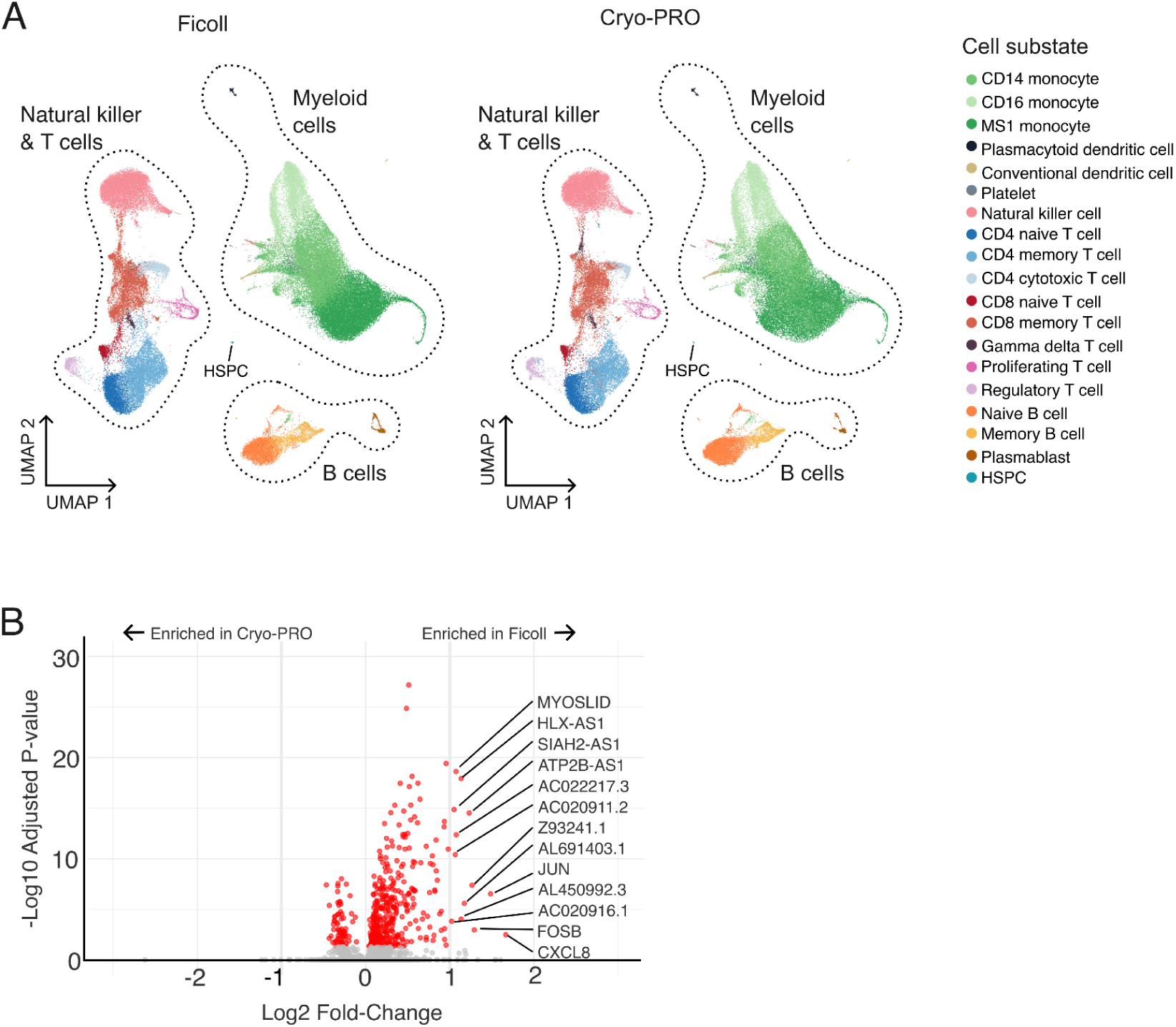
Comparison of cell substate identification and gene expression by method. (A) Two-dimensional uniform manifold approximation and projection (UMAP) of cells by processing method; Ficoll (left) and Cryo-PRO (right). Dotted outlines represent types of PBMCs. Cell substates were identified by clustering cells of each method independently; substate identities were then projected onto a shared set of UMAP axes (see Methods). (B) Volcano plot showing genes differentially up-regulated (positive log2FC) or down-regulated (negative log2FC) in Ficoll compared to Cryo-PRO using pseudobulk gene expression data from all cells (see Methods). Genes with adjusted p-values of less than 0.05 are shown in red; those with p < 0.05 and abs(log2FC) > 1 are labeled.

Top marker genes distinguishing each cluster were identified using the FindMarkers function in Seurat (20); rank was determined by fold-change of the gene expression within cells of each cluster compared to the cells outside of the cluster (24) (Supplemental Table 1). The average expression of key identifying genes (Figure 4, color scale) and cell surface proteins (Figure 5, color scale) was similar between Ficoll and Cryo-PRO methods for each cell type and substate, as was the proportion of cells for which these features could be detected (Figure 4-5, dot size). Our analysis identified the MS1 monocyte state previously discovered to be enriched in patients with sepsis (10, 25). Expression patterns for key MS1 marker genes were similar between Ficoll and Cryo-PRO samples (Supplemental Figure 3A).

**Figure 4.**
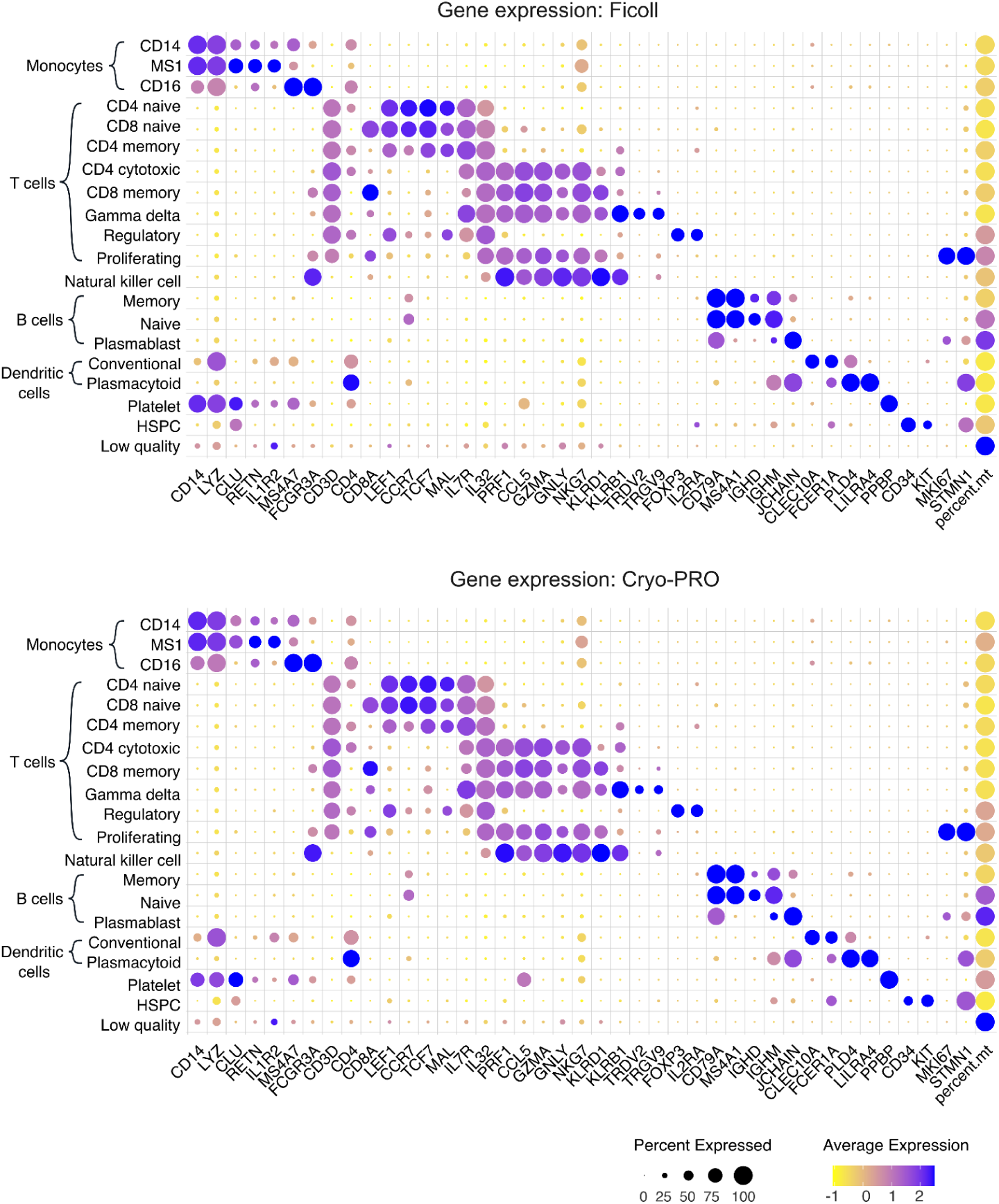
Dot plots of key marker genes and percent mitochondrial reads (percent.mt) for cell substates identified in scRNA-seq analysis by method (top: Ficoll, bottom: Cryo-PRO). Color represents scaled relative expression (blue = higher expression), and size represents the proportion of cells in each substate where the feature was detected.

**Figure 5.**
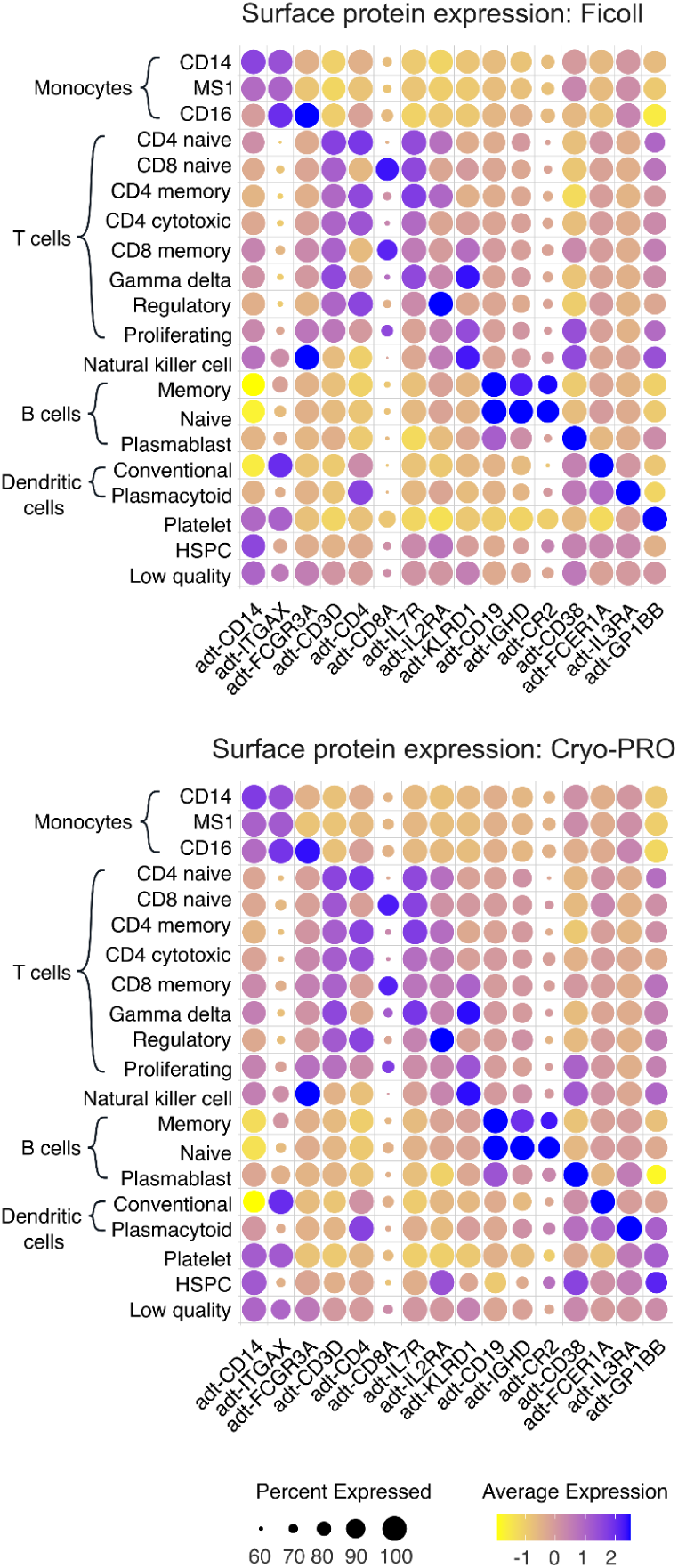
Dot plots of key surface marker proteins (detected using CITE-seq) for each cell substate. Color represents scaled relative expression (blue = higher expression), and size represents the proportion of cells in each substate where the feature was detected.

In an orthogonal approach, we used FindMarkers and the DESeq2 package in R to compare gene expression between all cells processed by the two methods to identify differentially expressed genes (Supplemental Table 2). Among statistically significant (p<0.05) genes, we did not observe substantial (greater than 4) fold-change differences in expression between the two methods. Most genes with more than a 2-fold expression change were non-coding genes; exceptions included CXCL8, FOSB, and JUN, which were slightly up-regulated in Ficoll samples (Figure 3B). Similar differentially-expressed genes were identified when comparisons were performed within each major cell type (Supplemental Figure 3B), rather than across all cells. Crucially, no genes used to identify cell types were differentially expressed by more than a 2-fold change. Pathway analysis could not be performed due to the sparse number of robustly differentially-expressed genes. However, immediate early genes are a class of genes commonly transiently upregulated in many types of cells as a primary response to a variety of stimuli; the presence of the immediate early genes JUN and FOSB may suggest an early response to *ex vivo* stimulation in Ficoll-processed cells (26, 27).

### Ficoll and Cryo-PRO yield similar immune cell type and substate abundances

Defining the composition of circulating immune cells and their substates on a per-patient level is an informative application of scRNA-seq. In sepsis, heterogeneity in the distribution of immune cell types and states between patients is thought to contribute to differences in illness trajectory, outcomes, and response to therapies (28). To evaluate agreement between methods for characterizing immune cell profiles in sepsis, we computed proportions for each cell type and substate for each sample processed using both Ficoll and Cryo-PRO methods. We defined proportion of cell type as the number of cells of a particular type (e.g., B cells) divided by the number of all PBMCs combined. For cell substates, proportion was defined as the number of cells assigned to a substate divided by the total number of cells of that cell type (e.g., number of CD16+ monocytes divided by the total number of monocytes). We then compared cell type proportions between paired Ficoll and Cryo-PRO samples from each of the subjects using correlations. Substate proportions were calculated for monocytes, T cells, B cells and dendritic cells; natural killer cells did not demonstrate unique substates, and substate proportions were therefore not computed.

Proportions of cell types were significantly correlated between methods (Figure 6A), with Pearson correlations (R) ranging between 0.87 and 0.95 (p<0.001 for all comparisons). For most substates, correlations were also positive and statistically significant (R values from 0.45 to 0.95; p<0.05); the exception being gamma delta T cells (R = 0.28, NS) (Figures 6B-D, Supplemental Figure 4). Gamma delta T cells were present at very low counts across methods, likely increasing the effects of outliers on correlations. Within monocytes, correlations across methods were higher for CD16+ monocytes (R = 0.93, p<0.001) than for CD14+ MS1 (R = 0.80, p<0.001) and classical CD14+ (R = 0.79, p<0.001). Comparison of cell substate proportions between Ficoll and Cryo-PRO at an individual subject level showed a high degree of similarity across each of the 24 subjects analyzed (Supplemental Figure 5, Supplemental Table 3), with especially close alignment for the 15 patients whose samples were processed immediately at clinical sites (Supplemental Figure 5A).

**Figure 6.**
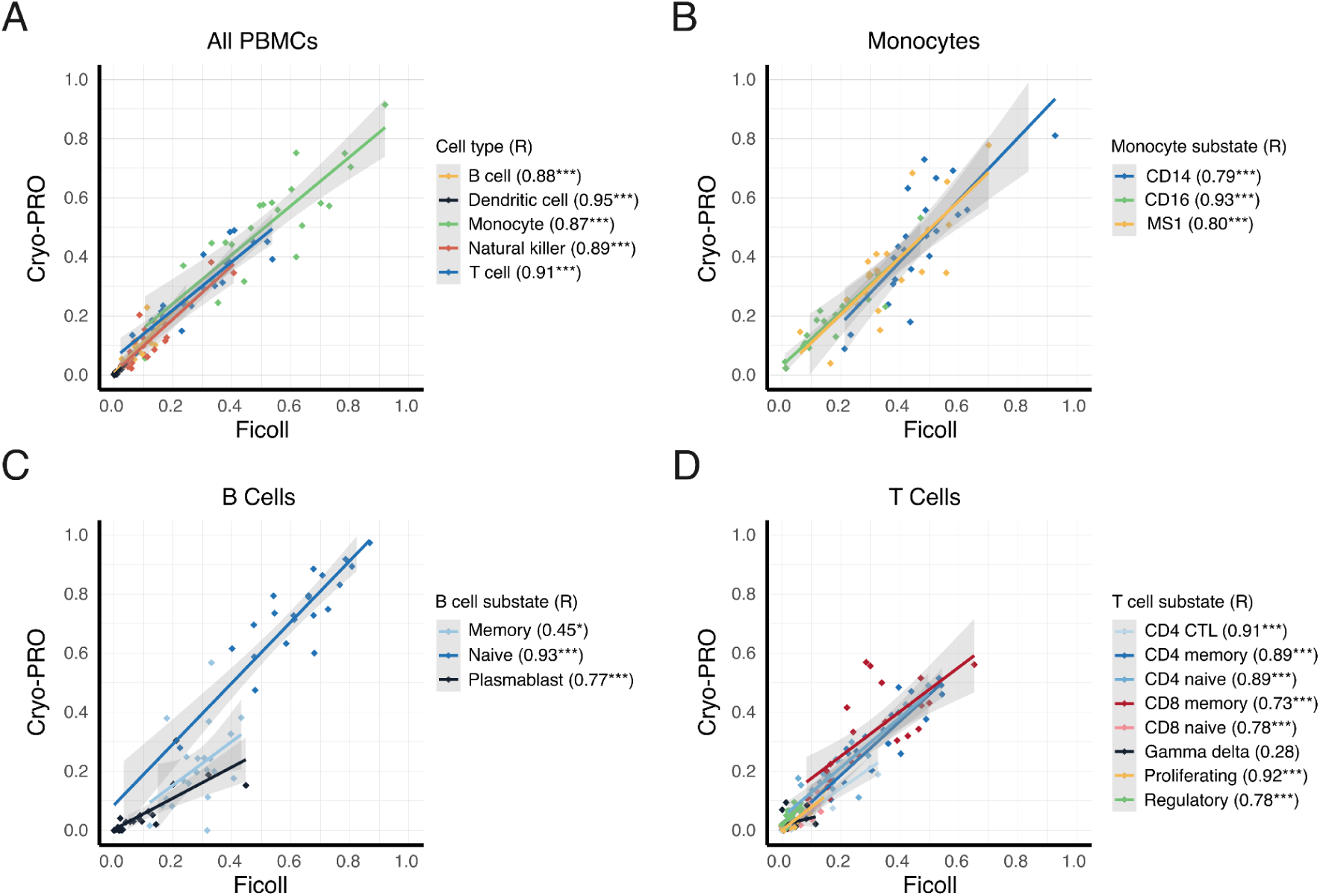
Correlation of cell type and substate proportions between Ficoll and Cryo-PRO methods. (A) Scatter plot of cell type proportion from Ficoll and Cryo-PRO. Each point represents the proportion of one cell type from one patient sample, as measured by each method. Each cell type is represented by a different color. Proportion is the number of cells of one cell type divided by the total number of PBMCs from that patient sample. Patient-paired Ficoll:Cryo-PRO samples are plotted to assess correlation by method for each patient. (B-D) Scatter plots of cell substate proportion from Ficoll and Cryo-PRO. Each point represents the proportion of one cell substate from one patient sample, as measured by each method. Each cell substate is represented by a different color. Proportion is the number of cells of one cell substate divided by the total number of cells from its cell type from that patient sample. Patient-paired Ficoll:Cryo-PRO samples are plotted to assess correlation in method for each patient. Trendlines indicate a linear regression fit with shaded 95% confidence intervals. Pearson’s correlations (R) are shown for all trendlines (*p < 0.05, ** p < 0.01, ***p < 0.001).

To evaluate the robustness of Cryo-PRO to processing location, we assessed the technical reproducibility of scRNA-seq results for the 8 patients whose samples were processed simultaneously at MGH and BIDMC, acknowledging inherent processing delays due to sample transport between sites (see Methods). Proportions of major cell types (monocytes, B cells, and T cells) were highly correlated between sites for Ficoll (R values from 0.83 to 0.96, p<0.001; Figure 7A) and for Cryo-PRO (R values from 0.86 to 0.99, p<0.001; Figure 7B). Dendritic cells (Ficoll R = 0.05, Cryo-PRO R = 0.44; both NS) and natural killer cells (Ficoll R = 0.34, Cryo-PRO R = 0.72; both NS) were poorly correlated between sites, possibly due to small overall cell counts and variable yield between processing runs that exaggerate differences in cell proportions. Correlations were significant for nearly all substates of monocytes, T cells, and B cells, and dendritic cells for each method between sites (Supplemental Figure 6), though for some substates including MS1, cross-site correlations were slightly lower for Cryo-PRO than Ficoll.

**Figure 7.**
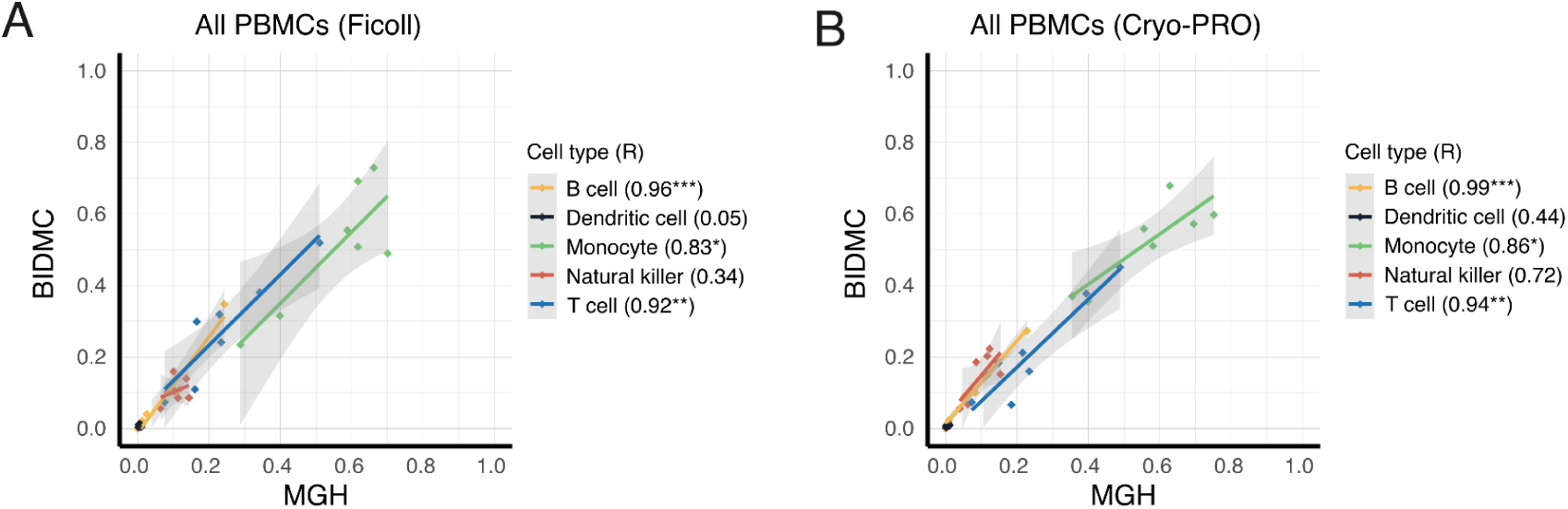
Between-site reproducibility of cell type proportion for samples processed at both locations, using Ficoll (A) or Cryo-PRO (B). Each point represents the proportion of one cell type from one patient sample, processed at each site. Each cell type is represented by a different color. Proportion is the number of cells of one cell type divided by the total number of PBMCs from that patient sample. The patient-paired Ficoll:Ficoll samples and Cryo-PRO:Cryo-PRO samples from the two different enrollment sites are plotted to assess between-site technical reproducibility. Trendlines indicate a linear regression fit with shaded 95% confidence intervals. Pearson’s correlations (R) are shown for all trendlines (*p < 0.05, ** p < 0.01, ***p < 0.001).

### Single-cell TCR sequencing yields comparable repertoire capture from Ficoll and Cryo-PRO samples

T cell lymphopenia and altered TCR diversity are recognized features of sepsis and its recovery. Previous studies have demonstrated reduced TCR repertoire breadth early in the course of septic shock (29, 30), with persistent contraction of the repertoire associated with increased mortality, higher rates of nosocomial infection (29), and reactivation of latent viral infections such as cytomegalovirus (30). These observations underscore the clinical importance of tracking TCR repertoire dynamics in sepsis. Capturing this data alongside paired single-cell gene expression data provides valuable insight into immune dysfunction within cellular substates, as well as gene expression programs associated with changes in clonotype diversity.

To determine whether paired single-cell transcriptomic and TCR profiling are preserved with Cryo-PRO, we applied the 10x Genomics 5′v2 Immune Profiling workflow (Methods) to matched patient samples processed using either Ficoll or Cryo-PRO. This strategy allows for joint recovery of full-length V(D)J sequences and gene expression data from the same cells. We compared the yield and quality of TCR sequencing across both methods.

Among Ficoll-processed samples, we recovered TCR sequences from 21,876 cells, representing 98.6% of T cells with transcriptomic data. For Cryo-PRO samples, we recovered TCR sequences from 18,447 cells (96.5% of T cells with transcriptomic data). Expanded and unique clonotypes were represented similarly on UMAP projections of Ficoll and Cryo-PRO T cells (Figure 8). As expected, effector and cytotoxic cells (CD4+ cytotoxic T cells and CD8+ memory T cells), which expand during acute immune responses to infection (31, 32), displayed the highest levels of clonal expansion by both methods, whereas naïve CD4+ and CD8+ T cell substates displayed greater sequence diversity and fewer expanded clonotypes (Figure 8).

**Figure 8.**
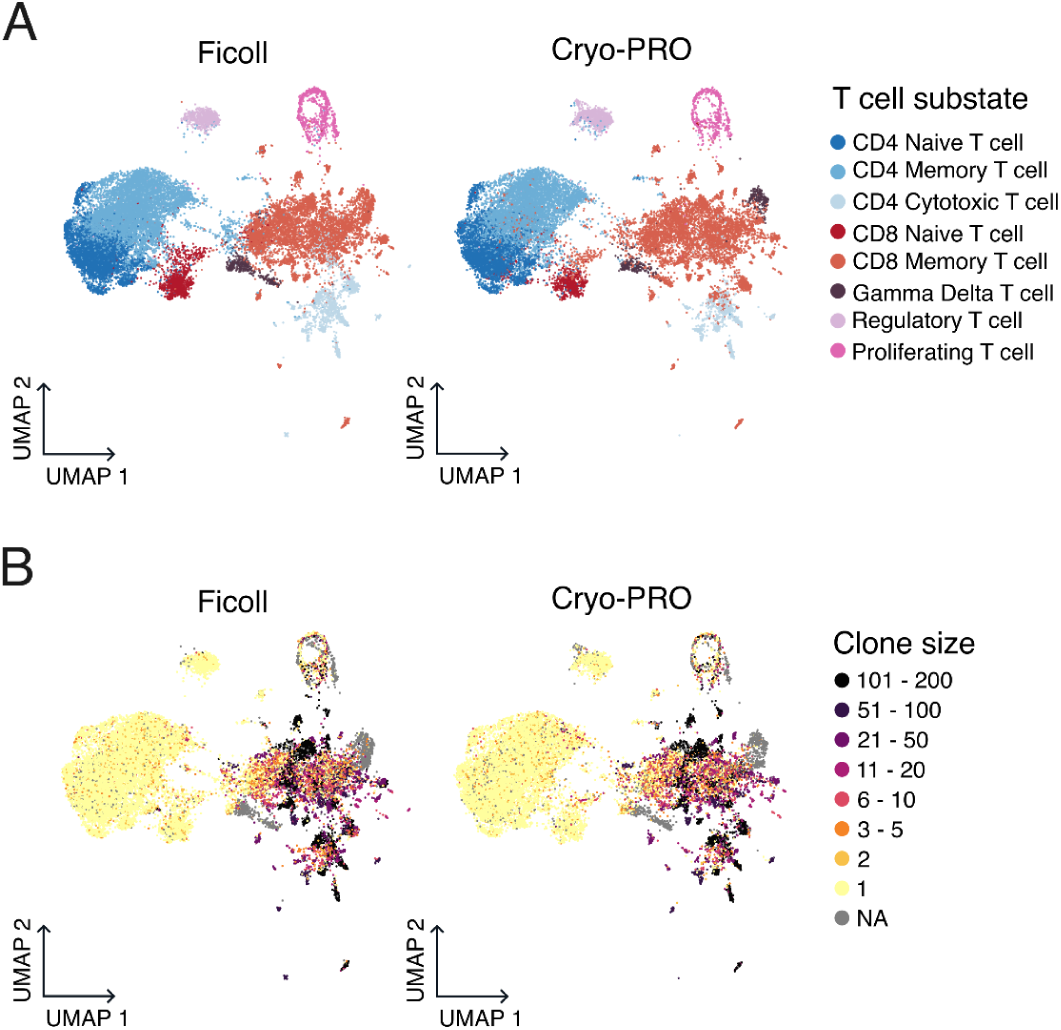
T cell substates and corresponding TCR repertoire clonality by method. UMAP of T cells profiled, split by method (left: Ficoll, right: Cryo-PRO). Cells are colored by (A) their cell substate, or by (B) the number of identical TCR sequences to represent clone size (darker = greater clonal expansion).

Per-patient clonotypes were generally similar between methods in the proportion of expanded clones (Supplemental Figure 7). Discrepancies in clonal proportions were generally attributable to low T cell recovery from at least one of the samples (Supplemental Figure 7). For patients with samples processed at both clinical sites, we observed similar trends, with clonal proportions closely resembling each other between processing center and method for each patient profiled (Supplemental Figure 7).

We next compared exact nucleotide sequences of the captured TCR clonotype transcripts between methods. Our expectation was that while many clonotypes could be present at very low frequencies in a given sample, expanded clonotypes would be detected at higher frequencies in both Ficoll and Cryo-PRO samples from the same patient. For each patient, we calculated the abundance of each matching unique TCR sequence as a proportion of the total TCR sequences and compared these proportions between methods and processing centers. Patient-matched Ficoll and Cryo-PRO TCR sequence proportions were positively correlated (Pearson’s R = 0.47, p<0.001). Significant overlap was also observed between processing centers for the eight patients processed at MGH and BIDMC, with higher cross-site reproducibility for Cryo-PRO (R = 0.79, p<0.001) than for Ficoll (R = 0.38, p<0.001) (Supplemental Figure 8). On a per-patient basis, method and processing center comparisons showed similar patterns, with differences likely reflecting variable T cell recovery (Supplemental Figures 9,10).

### Ficoll and Cryo-PRO methods preserve cellular functions required for phagocytosis

Finally, we sought to measure functional activity in our cryopreserved cells. A key monocyte function is phagocytosis, which requires cells to detect the presence of a pathogen, encapsulate it inside a phagosome, and initiate microbial killing and degradation via fusion with lysosomes and subsequent exposure to hydrolytic enzymes and acidic conditions (33). Detection of phagocytosis therefore indicates coordination of cellular signaling pathways, cytoskeletal rearrangement, and functional organelles. We measured phagocytic activity in PBMCs from samples processed using Ficoll and Cryo-PRO as a means of assessing cell viability, function, and responsiveness to environmental stimuli.

Ficoll and Cryo-PRO samples (one of each from nine sepsis patients and one healthy subject) were collected, preserved, and frozen. Both sample types then underwent all of the processing steps before flow cytometry sorting, including the red blood cell depletion step for Cryo-PRO samples only (Methods). Cell suspensions were incubated with *E. coli* pHrodo Bioparticles (Invitrogen), which fluoresce in acidic phagolysosomes (34). After incubation, cells were fluorescently stained for viability, CD45, CD15, and CD14, and analyzed with flow cytometry.

We measured phagocytic activity as the mean fluorescence intensity (MFI) of the pHrodo Bioparticles, which emit light at green (FITC) wavelengths. We first confirmed that live PBMCs (DAPI-CD45+ CD15-cells) had greater pHrodo MFI upon addition of the bioparticles (Supplemental Table 4), consistent with engulfment into phagolysosomes. We next stratified the live PBMCs by CD14 expression. Assessment of control wells lacking bioparticles confirmed that fluorescent spillover from the CD14 stain did not impact the pHrodo MFI measurement (Supplemental Table 4). CD14+ monocytes are the most abundant phagocytes within PBMCs (35), and within our samples, were expected to show higher pHrodo MFI compared to the CD14-fraction, which consists primarily of lymphocytes with low phagocytic activity, but also includes CD16+ monocytes and dendritic cells (36). Accordingly, across all individuals, CD14+ PBMCs exhibited higher phagocytic signal than CD14-PBMCs using either processing method (median pHrodo MFI fold-change of ∼3.98 for Ficoll and ∼4.72 for Cryo-PRO), despite substantial inter-individual variability (Figure 9a). The pHrodo MFI of CD14+ cells was generally higher in Ficoll than in the corresponding Cryo-PRO sample (median of 30,240 RFU across samples compared to 4,946 RFU across samples) (Figure 9b).

**Figure 9.**
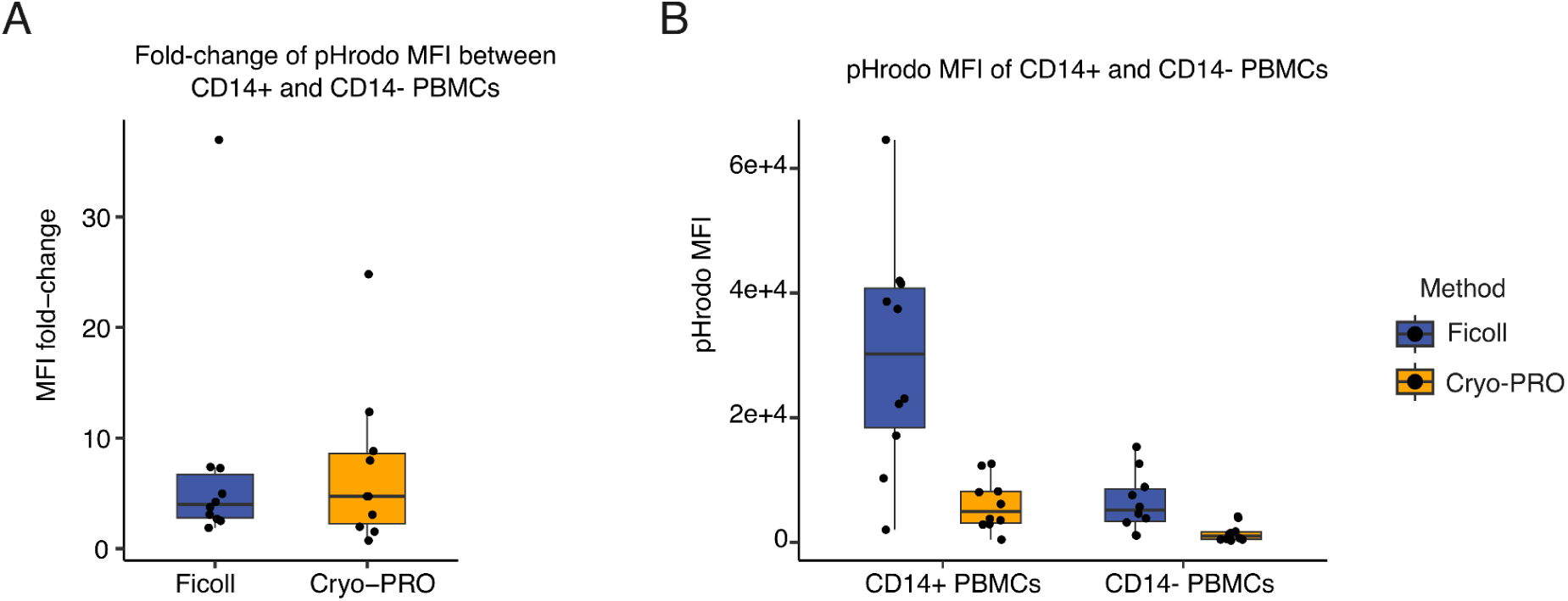
Cryo-PRO preserves phagocytic function in peripheral blood mononuclear cells (PBMCs). (A) Fold-change differences of pHrodo mean fluorescence intensity (MFI) between CD14+ and CD14-PBMCs for Ficoll (blue) and Cryo-PRO (yellow) samples. (B) Absolute pHrodo MFI of PBMCs, stratified by CD14+ expression (horizontal axis) and method (blue = Ficoll, yellow = Cryo-PRO). Boxplots indicate the median and IQR (whiskers = 1.5x IQR). Points represent individual samples.

This difference may reflect competition for bioparticles during the incubation before flow cytometry, due to greater carryover of additional phagocytes (e.g., granulocytes) in Cryo-PRO suspensions; such competition would reduce the effective particle-to-CD14+ cell ratio even though CD15+ granulocytes are subsequently excluded from analysis by gating. Accordingly, direct between-method comparisons of CD14+ pHrodo MFI are confounded due to differences in cellular composition during incubation, whereas the within-method contrast (CD14+ vs CD14-) demonstrates that preserved CD14+ PBMCs retain detectable hallmark phagocytic function with both methods, with comparable increases in bead uptake over CD14-cells.

## DISCUSSION

Single-cell transcriptional profiling facilitates high-resolution characterization of the heterogeneity among circulating immune cells, thereby revealing critical insights into diseases such as sepsis where the immune response plays a pivotal role (37). Performing these investigations with clinical samples is critical for translational goals, as it establishes a direct link between cellular transcriptomics and patient-derived data. However, the current state-of-the-art process for single-cell profiling faces a number of major roadblocks to application on clinical samples: intensive sample collection strategies that require more time, equipment, and molecular techniques than are typically available to clinical study teams; and cost. Although scRNA-seq is becoming more economical with emerging technologies and the ability to pool samples, performing scRNA-seq from patient blood still requires PBMC isolation via the time-and resource-intensive process of Ficoll density gradient centrifugation. This limitation has greatly constrained the application of scRNA-seq in clinical investigations, resulting in smaller clinical cohorts that may not fully capture the heterogeneity of diseases under study.

Here, we demonstrate that direct cryopreservation of a small volume (∼1 mL) of whole blood at the point of care, followed by thawing and PBMC isolation at a centralized research facility, is a viable alternative to on-site Ficoll processing for scRNA-seq, CITE-seq, and TCR repertoire profiling. This simple and streamlined approach significantly reduced the time and technical expertise needed to obtain clinical samples, while still preserving single-cell transcriptomes and surface proteomes in patients with sepsis. We independently identified the same immune cell types and substates in the datasets of Cryo-PRO and Ficoll, including the sepsis-enriched monocyte substate MS1, considered important in sepsis immunopathophysiology (10, 38). We found a high correlation between methods for the abundances of all major cell types. Although cell substates may be less distinctly defined by their transcriptional profile than cell types and are therefore more susceptible to misidentification due to stochasticity in clustering, we still observed high correlations between most substate proportions derived from the two methods after independent clustering and substate assignment. Moreover, patterns of gene and surface protein expression were similar across cell types and substates, with minimal differential gene expression between methods. TCR capture from T cells processed with Cryo-PRO yielded sequences and proportions of expanded clonotypes similar to those of Ficoll. Lastly, phagocytosis assays demonstrated preservation of this innate immune cell function across CD14+ cells purified by either Ficoll or Cryo-PRO. Together, this substantial equivalence between the gold-standard method of Ficoll processing and Cryo-PRO demonstrates that Cryo-PRO does not introduce major artifacts from processing and generates biologically meaningful results in patients. When deployed across two different enrolling emergency departments, cell type and substate abundances from Cryo-PRO showed strong correlations across sites. This finding shows that Cryo-PRO is robust to variations in collection site and operator, further supporting it as a reliable strategy for expanding multimodal single-cell immune profiling studies.

Other forms of rapid whole blood cryopreservation have recently been shown to be compatible with scRNA-seq (39, 40). In one of these studies, a substantial loss in the fractional abundance of myeloid cells was observed when compared with samples obtained using Ficoll (40). Our approach produces better equivalence with the standard Ficoll method across immune cell types. Another method (39) is based on the use of fixed cells, which provides more flexibility in the cryopreservation process compared to Ficoll (41). However, because fixation impairs polymerases involved in cDNA library preparation, fixed cell RNA profiling requires hybridization to a predefined set of probes, rather than sequencing, to detect transcripts (42), introducing a number of limitations. In particular, hybridization-based approaches require *a priori* knowledge of the cell’s potential transcriptional signature, and thus fail to capture regions of high allelic diversity such as TCR (and B cell receptor) clonotypes.

Other options for rapid sample processing and preservation with fixed cell profiling are generally kit-specific (43), requiring users to commit to an approach prior to the start of sample collection, use costly reagents for all collected samples, and sacrifice the potential for diverse allele region capture before experimentation even begins. Cryo-PRO enables kit-agnostic preservation that simplifies and expedites sample collection, preserving the option to later profile diverse allele regions such as TCR clonotypes. Lastly, fixed cell profiling necessitates killing the cells. We found that Cryo-PRO preserves cells capable of phagocytosis, suggesting that cells retain their phenotype and are still responsive to environmental stimuli. Many sequencing studies require such active cellular functions, for example, measuring cells’ transcriptional responses to stimuli (44) or Perturb-seq (45). In addition, some of the aforementioned approaches, in part due to smaller sample size, relied on co-clustering of scRNA-seq data with the traditional Ficoll method to assign cell states (39, 40). In order to be useful at the point of care, a streamlined collection method must stand alone; we therefore independently clustered and analyzed patient-matched data from Cryo-PRO and Ficoll, and observed high technical and biological equivalence.

The Cryo-PRO method has transformative potential for multicenter sample collection and clinical trial enrollment efforts by greatly simplifying on-site protocols for scRNA-seq and transferring the technically demanding steps to a centralized location. The resource demands of onsite processing for scRNA-seq particularly impede studies of highly heterogeneous diseases with acute onset where study collection strategies must be deployable at any time a patient may present. Sepsis is an archetype of such a condition, and sample sizes for scRNA-seq studies of sepsis have, as a consequence, been underpowered to bring the full power of the method to bear on investigating biological reasons underlying the clinical heterogeneity of the condition (10, 12–15). The substantial reduction in technical skill and time requirements for sample processing and preservation (i.e., mean time of 13 minutes for Cryo-PRO vs 143 minutes for Ficoll) has crucial operational implications in the clinical research setting. Simplifying sample collection also offers an opportunity for improving cost efficiency by enabling the rapid enrollment of many potentially suitable patients for clinical studies, followed by retrospective adjudication to inform the selection of appropriate patients for sequencing. Widening the net of subjects enrolled in this manner would better reflect the true patient heterogeneity in conditions under study, enabling post-hoc enrichment for rare clinical phenotypes or outcomes. For sepsis, such a strategy could facilitate the derivation of scRNA-seq-based endotypes across large, diverse cohorts, including those from hospitals in underserved communities without dedicated research teams and resources to typically participate in clinical research.

Our study has several limitations. First, our cohort size remains small in the context of sepsis studies. However, to the best of our knowledge, this study (n = 24 subjects and 32 paired samples) is the largest to date evaluating the feasibility of whole blood cryopreservation for scRNA-seq, CITE-seq, and TCR profiling in any context, and demonstrates substantial equivalence with conventional methods. We aim to further validate Cryo-PRO as a sample processing approach in a larger cohort of subjects in the future. Second, although all major cell types and substates had substantial equivalence in patient-level abundance, some cell substate abundances differed between Cryo-PRO and Ficoll methods. Some differences in substate assignment within cell types (e.g., MS1 versus classical CD14+ monocytes or memory versus naive B cells) are less well-defined than differences in cell types, and may reflect more of a continuum than a dichotomy, so more stochastic differences in assignments are expected. Other cell substates like gamma delta T cells and substates of dendritic cells were present at very low abundances and therefore more susceptible to outlier effects. While some differences between methods may reflect differences in either gene expression or survival by cell type and substate, each method introduces processing steps that may perturb transcription, i.e., centrifugation for 2 hours through a density gradient followed by freezing, thawing, and flow cytometry for Ficoll; or exposure to DMSO, freezing, thawing, magnetic cell separation, and flow cytometry for Cryo-PRO. Additionally, 8 subjects were processed in parallel at two sites, with a resulting mean delay of just over 2 hours prior to processing by either method, which may have affected subsequent sequencing results in these samples. Despite these limitations, the overall agreement in cell type and substate proportions between methods, and the minimal perturbations seen in differential expression by method, suggest that major transcriptional signals that reflect relevant biology are preserved. Third, we evaluated one function (phagocytosis) of the PBMCs isolated by Cryo-PRO; additional assays will be needed to further assess the functional capacity of PBMCs isolated using the Cryo-PRO method..

By simplifying sample collection at the point of care, Cryo-PRO can unlock greater potential of scRNA-seq to study the biology of complex clinical conditions across multiple collection sites, including lower-resource settings, thus improving capture of the true heterogeneity of diseases. This method greatly lowers the barrier to embedding scRNA-seq-compatible collection strategies in randomized clinical trials, which would enable post-hoc analyses to identify biological subsets of patients (i.e., endotypes) that may selectively respond to therapeutic interventions. In addition, Cryo-PRO could enhance the cost-efficiency of scRNA-seq by enabling “overcollection” at the point of care, reserving PBMC isolation and scRNA-seq only for samples from patients who display clinical phenotypes or disease trajectories of interest on subsequent adjudication. Thus, Cryo-PRO has substantial potential to expand the application of scRNA-seq towards personalized medicine in complex and heterogeneous conditions like sepsis, and this work represents an important step towards that goal.

## METHODS

### Sex as a biological variable

We included 8 female patients and 15 male patients in our study, in addition to one male healthy control subject. To account for patient heterogeneity in our study, including potential effects of sex as a biological variable, all comparisons in our study were made by comparing samples processed with different methods but collected from the same subject.

### Patient enrollment and clinical adjudication

This study was conducted at Massachusetts General Hospital and Beth Israel Deaconess Medical Center. Inclusion criteria were adult patients arriving at the Emergency Department with evidence of organ dysfunction for whom bacterial infection was possible or suspected. Eligible patients had a blood sample collected under an IRB-approved alteration of informed consent, which allowed a research sample to be drawn simultaneously with the initial clinical blood draw. Informed consent was obtained from the patient or a surrogate at a later time after initial resuscitation.

Samples were collected for 100 patients during a 12-month period from April 2023 to March 2024. Of these, 84 provided consent for research use and were considered enrolled; samples from patients who did not provide consent were discarded. Clinical data were collected on all enrolled subjects and entered into REDCap by clinical research coordinators. Physician adjudication (MRF) was later performed via retrospective chart review with access to all available clinical data and notes during the subject’s hospitalization. Subjects were adjudicated as meeting Sepsis-3 criteria (18) for sepsis or septic shock during the first 48 hours of hospitalization, or as having infection without sepsis versus a non-infectious cause of illness. For the current analysis, we prioritized sequencing in those subjects adjudicated as sepsis and septic shock, and selected 23 for sequencing.

### Sample collection at clinical sites

Research blood samples were collected in 10-mL EDTA tubes. Up to 20 mL was collected if patient samples were being parallel-processed at both clinical sites; up to 10 mL was collected if patient samples were being processed at only a single site. For samples parallel-processed at both clinical sites, one of the two 10-mL tubes collected at the enrolling site was couriered to the second site.This resulted in a delay in processing of about 2 hours on average; samples from one subject were delayed >3 hours. Samples obtained for single-site processing were taken directly to the onsite lab for immediate processing. All 10-mL samples underwent cryopreservation of whole blood (2 mL) and onsite density gradient centrifugation with Ficoll (∼3 to 6 mL) as described below. Up to 3 mL of the collected whole blood sample was used for other research purposes.

### Onsite whole blood cryopreservation for Cryo-PRO

For immediate whole blood cryopreservation, 2 mL of blood was mixed with 200 uL DMSO (final 10% v/v). Two 1-mL aliquots in cryovials were then prepared per sample and were slowly cooled using a Mr Frosty (Sigma-Aldrich) at -80 °C. Aliquots were stored onsite at -80°C for less than 1 month before being transported to the Broad Institute (Cambridge, MA) on dry ice and immediately stored at -140°C until the time of sequencing. Duplicate aliquots served as backups.

### Onsite density gradient centrifugation (Ficoll) and cryopreservation of PBMCs

Density gradient centrifugation was performed on the remaining blood in the EDTA tubes after cryopreservation aliquots had been obtained (∼3 to 6 mL). Blood with EDTA was diluted 1:1 with room temperature PBS and layered over Ficoll-Paque PLUS density gradient media (Cytivia) in SepMate tubes (STEMCELL), then centrifuged at 1,200 rcf for 20 minutes at 20°C with slow acceleration and the brake off. The buffy coat layer was collected and washed twice with cold RPMI (Gibco) before cells were counted, resuspended in CryoStor CS10 (STEMCELL), and aliquoted into cryotubes targeting 1 million cells per vial. Samples were cooled, stored, and transported in the same manner as the Cryo-PRO samples.

### Healthy donor blood cryostorage

Fresh healthy donor blood in EDTA tubes was obtained from Research Blood Components (Watertown, MA) and processed within two hours of receipt. Whole blood Cryo-PRO and Ficoll PBMC cryostorage steps were performed as above, though all processing steps occurred at the Broad Institute.

### Pre-sequencing processing for both Ficoll and Cryo-PRO samples

All subsequent processing and analysis steps were performed at the Broad Institute. On the day of flow cytometry sorting and Chromium 10X processing, a sample of cryopreserved whole blood (for Cryo-PRO) and a Ficoll sample were thawed for each patient. Sequencing batches were designed to contain four Ficoll samples and four patient-matched Cryo-PRO samples to minimize the effect of sequencing batch variation on the method comparison; therefore, 8 samples total were processed in parallel.

For each of the four Cryo-PRO samples, 1 mL of cryopreserved whole blood was thawed in a 37°C water bath for 1 min 15 seconds and transferred into a 5-mL polystyrene round-bottom tube using 1 mL of PBS containing 2 mM EDTA and 2.5% FBS. Samples were immediately depleted of red blood cells using the STEMCELL EasySep RBC depletion kit. Briefly, the diluted blood was mixed with 50 uL of the RBC depletion reagent before immediately being placed on a magnet for 5 minutes at room temperature. The supernatant was pipetted off and mixed with an additional 50 uL of RBC depletion reagent in a new tube before another immediate 5 minute magnet incubation. At the end of the second incubation, the supernatant was transferred into 8.5 mL of FBS-RPMI (RPMI + 10% FBS + 1x penicillin/streptomycin) on ice. These steps were performed in parallel for the four Cryo-PRO samples.

For each of the four Ficoll samples, one vial per patient was thawed in a 37°C water bath for 1 min 15 seconds before transfer with 1 mL of FBS-RPMI into 8.5 mL of FBS-RPMI on ice. For patients with three or more Ficoll vials, two vials were thawed and combined to improve cell recovery. These steps were performed in parallel for the four Ficoll samples.

For the subsequent steps, Cryo-PRO and Ficoll samples were processed identically and in parallel. All samples were centrifuged to pellet the cells (300xg, 5 minutes, 4°C), then resuspended with FACS-PBS (PBS + 2mM EDTA + 2.5% FBS) and centrifuged again. Each sample was then resuspended in 50 uL FACS-PBS and incubated on ice with a hashtag oligo for pooled sequencing (TotalSeq™ anti-human Hashtags, BioLegend), an Fc receptor blocking solution (Human TruStain FcX™, BioLegend), and flow cytometry stains (DAPI solution,Thermo Scientific; Alexa Fluor® 700 anti-human CD15 [Clone: HI98], BioLegend; FITC anti-human CD235a [Clone: HI264], BioLegend; and PE anti-human CD45 [Clone: HI30], BioLegend). Samples were then washed in cold FACS-PBS and sorted on a SONY MA800 cell sorter to select DAPI-CD15-CD235a-CD45+ cells, targeting 50,000 cells per sample.

After sorting, the hashed and sorted cells from all eight samples were pooled, pelleted (300xg, 5 minutes, 4°C in FACS-PBS), and resuspended in a CITE-Seq cocktail for surface proteome measurement (19) for a final incubation on ice. After 20 minutes, the cells were washed twice more (centrifugation at 300xg, 5 minutes, 4°C followed by resuspension in PBS + 2.5% FBS), counted, and resuspended in PBS + 2.5% FBS for a target concentration of 1,000 cells/uL.

### Library construction and scRNA sequencing

Droplet-based single-cell RNA capture and RNA and ADT library construction was performed with the Chromium single-cell 5’ kit v2 (10x Genomics, Inc). Forty uL of cells were loaded onto the Chromium Chip K, and Gel Bead-in Emulsion creation and library construction followed according to the manufacturer’s protocol (46). Eight batches of libraries were prepared (including gene expression libraries and cell surface protein libraries), with each batch barcoded using the 10X Dual Index Kit and sequenced altogether. Gene expression libraries were initially sequenced at low depth (∼200 reads/cell) using the Illumina MiniSeq 150 Cycle Hi-Output Kit for a quality check and cell count estimate to inform library balancing. Rebalanced libraries targeting 50,000 reads/cell for gene expression and 10,000 reads/cell for surface proteins were then sequenced on an Illumina NovaSeq S4.

### Data preprocessing

FASTQ files were aligned to a reference genome (GRCh38) using the Cell Ranger v6 pipeline by 10X Genomics. Demultiplexing and multiplet detection with patient hashtag oligos was performed using the Cumulus pipeline (47). Filtered gene expression matrices and CITE-Seq matrices were then analyzed using the Seurat V5 package in R. Multiplets and cell barcodes without corresponding gene expression, CITE-seq, and demultiplexing data were removed. Genes present in less than 10 cells were removed. Sequencing data from each method was split into two datasets and analyzed independently. For each set, RNA expression data was normalized, scaled, and integrated between sequencing batches using the top 2,000 most variable genes. Scaled CITE-seq data was integrated by finding multimodal neighbors using the first 50 principal components of RNA and CITE-seq data.

### Clustering and substate identification

Clustering was performed using the resulting weighted-nearest-neighbors graph, and the Clustree package (48) was used to determine clustering resolution. Cell types were assigned to clusters using top marker genes for each cluster (determined by Wilcoxon rank-sum test, Bonferroni-corrected p-value < 0.05, ranked by fold-change), and cell substates were assigned using top marker genes obtained by subsetting and re-clustering cells from each cell type at a higher resolution. Classification of cell types and substates was cross-referenced using the annotated Azimuth reference dataset (20). Clusters were defined as low quality if over 20% of cells in the cluster were cells with mitochondrial genes representing 10% or more of total genes detected in that cell. Low quality clusters were removed from further analysis as part of an extended quality control. After method-independent cell substate assignment, the Ficoll and Cryo-PRO datasets were combined and a UMAP was generated using the weighted-nearest-neighbors graph for the purpose of data visualization.

### Differential gene expression

Statistical analysis of differentially expressed genes was performed using the DESeq2 package as described below (Statistics).

### Cell type and substate abundance

PBMC cell type proportions were calculated as a fraction of all major cell types (monocytes, B cells, T cells, NK cells, and dendritic cells). Cell substate proportions were calculated as a fraction within the parent cell type. Cell clusters defined as low quality, or belonging to a class of cells other than PBMCs (e.g., platelets and hematopoietic stem and progenitor cells), were not included in proportion calculations. Samples with fewer than 1,000 total cells were not included in correlation calculations (Figure 6-7; Supplemental Figures 4,6). When possible, the Ficoll:Cryo-PRO comparisons were made using samples processed at the same site at which they were collected. In the case of Subject 17, one sample yielded less than 1,000 cells, so the Ficoll:Cryo-PRO samples processed at the alternative site were used in calculations instead. Pearson correlations (R) were calculated and reported for scatterplots of cell types and substates.

### TCR sequencing and repertoire analysis

Paired α/β TCR sequences were obtained using the 10x Genomics 5′ V(D)J Immune Profiling workflow (). Following single-cell capture and cDNA amplification, TCR libraries were constructed in parallel with gene expression libraries from the same droplets, according to the manufacturer’s protocol. Libraries were sequenced to sufficient depth to recover full-length V(D)J transcripts. TCR reads were processed using Cell Ranger pipelines to assemble CDR3 sequences for both TCRα and TCRβ chains. Cells lacking a productive TCR sequence were excluded. Productive paired TCRαβ chains were extracted using the combineTCR() function in the scRepertoire R package (49). Clonotypes were defined by identical CDR3 amino acid sequences for both α and β chains, and clonal expansion was visualized using scRepertoire functions.

### Phagocytosis assays

Samples were collected and stored as described above. One Ficoll and one Cryo-PRO sample from ten subjects (nine patients, one healthy subject) were used. Due to limited samples per patient, technical replicates were not possible. Sample thaw followed steps in pre-sequencing processing through the first wash in FACS-PBS. Each sample was then split into two; one aliquot of each sample was incubated with reconstituted pHrodo Green *E. coli* BioParticles Conjugate for Phagocytosis (Invitrogen) for 10 minutes at 37 C; the other aliquot underwent identical handling without particles (control). After 10 minutes, samples were washed according to manufacturer protocol and stained for flow cytometry. Cells were stained using APC anti-human CD15 [Clone: HI98], Spark PLUS UV395™ anti-human CD14 [Clone: S18004B], PE anti-human CD45 [Clone: HI30] (BioLegend), and DAPI (Invitrogen), and then incubated on ice for 20 minutes. Cells were then washed in cold FACS-PBS, resuspended in 200 uL of FACS-PBS, and analyzed for surface markers using a Beckman CytoFLEX LX flow cytometer. Phagocytosis was measured simultaneously using the FITC channel. Data were analyzed using FlowJo to identify and stratify cell populations of interest (CD14+ PBMCs and CD14-PBMCs), and to quantify phagocytosis using mean FITC fluorescence intensity. Potential confounding effects of spillover, while measured to be minimal, were accounted for using a compensation matrix at the time of acquisition. Additionally, all reported FITC MFI values of cell populations with bioparticles were normalized by subtracting the FITC MFI of the corresponding population in the no-particle control wells to account for spillover into the FITC channel.

### Figure generation

Figure 1 was created in BioRender (50). Subsequent figures were generated in R using the ggplot2 package, the ScCustomize package (51), the Seurat package (20), and the scRepertoire package (49).

### Statistics

To assess differential gene expression between methods, scRNA-seq data was first pseudo-bulked by sample (generating 32 “bulk” RNA-seq samples from each method) to minimize p-value inflation (52), and FindMarkers with the DESeq2 package was used to detect differentially expressed genes. The same approach was applied within cell types by pseudo-bulking cells by sample x cell type. Unless otherwise stated, tests were two-sided. For differential gene expression analyses, p values were adjusted for multiple testing using the Benjamini–Hochberg false discovery rate; we report adjusted p values. For marker-gene discovery, genes were included if log2 fold-change >0.25, the genes were expressed in over 25% of cells in the cluster; and a Bonferroni-corrected p-value <0.05. For correlation analyses, we report Pearson’s R, with statistical significance assessed via two-sided t test. Where significance annotations are shown, thresholds were: p<0.05 (*), p<0.01 (**), p<0.001 (***).

### Study approval

This study was approved by the Massachusetts General Brigham IRB (2022P002833). Research blood was collected under an IRB-approved alteration of informed consent; written informed consent was subsequently obtained from participants or legally authorized representatives.

### Data availability

scRNA-seq data will be deposited to the Broad Institute Single Cell Portal (https://singlecell.broadinstitute.org/single_cell) on publication. Code used for data analysis will be available at github.com/alyssa_dubois/CryoPRO.

## ACKNOWLEDGMENTS

This work was supported in part by the National Institutes of Health (award nos. NIH R21GM148826 and NIH R01AI153142). The content is solely the responsibility of the authors and does not necessarily represent the official views of the NIH. The Broad Institute and Massachusetts General Hospital co-filed a provisional patent application on the disclosed subject matter; AKD, POA, MRF, and RPB are co-inventors of said application. We thank members of the patient enrollment team: Justin Margolin, Austin Vyas, Antoinette Nelson-Rodriguez, Kimberly Redman, Anna Grafals, Jodens Didie, and Carlo Ottanelli. We thank Miguel Reyes, Kiana Billman, Abraham Sonny, Nir Hacohen, Marcia Goldberg, Paul Blainey, Kamil Slowikowski, and Christopher Cosgriff for valuable discussion and input on methods and analysis, Leslie Gaffney for input on figure preparation, and the Broad Institute Technology Space (especially Andrew Patentreger) for support with sample processing.

## AUTHOR CONTRIBUTIONS

MRF, NIS, and RPB conceived the study.

MRF and NIS supervised enrollment and sample collection and performed clinical adjudications.

ACC, RH, OKN, CAZ, MKD, SM, AM, and BAP consented and enrolled patients and performed onsite processing steps.

MRF, RPB, POA and AKD designed the experiments.

AKD conducted experiments.

AKD and POA performed data analysis.

AKD, POA, MRF, and RPB prepared the manuscript.

AKD and POA contributed equally to the manuscript; authorship order was assigned by duration of involvement in the project.

---

**Supplemental Figure 1.** Number of singlet cells sequenced per method. Starting blood sample volume was variable for Ficoll samples and fixed at 1 mL for Cryo-PRO samples.

**Supplemental Figure 2.** Per-sample distributions of **(A)** UMIs of RNA transcripts, **(B)** unique genes, **(C)** percentage of mitochondrial transcripts, **(D)** unique surface protein features detected via CITE-seq, and **(E)** UMIs of surface protein features detected via CITE-seq per cell. Batches represent samples that were thawed, processed and sequenced together. Ficoll and Cryo-PRO samples from the same patient are plotted next to each other. For patients where parallel processing occurred at both clinical sites (bottom rows), the samples processed at the opposite site of enrollment are shown in lighter shades. A total of 137 different surface proteins were queried in the CITE-seq analysis. Violin plots show the distribution of each metric detected per cell; embedded boxplots indicate the median and IQR (whiskers = 1.5x IQR). PRO denotes Cryo-PRO.

**Supplemental Figure 3. (A)** Dot plots of marker gene expression by each monocyte substate. Color represents scaled relative expression (blue = higher expression). Size represents the proportion of cells in each substate where the feature was detected. **(B)** Volcano plots showing genes differentially up-regulated (positive log_2_FC) or down-regulated (negative log_2_FC) in Ficoll compared to Cryo-PRO after pseudobulk analysis. Genes with adjusted p-values of less than 0.05 are shown in red; those with p < 0.05 and abs(log_2_FC) > 1 are labeled. Plots are shown for differential gene expression among all cells (top left) and for each major cell type (subsequent plots).

**Supplemental Figure 4.** Scatter plot of dendritic cell substate proportions (i.e., the number of cells of a cell substate divided by the total number of dendritic cells from that patient sample) from Ficoll and Cryo-PRO. Each point represents the proportion of one cell substate from one patient sample, as measured by each method. Each cell substate is represented by a different color. Patient-paired Ficoll:Cryo-PRO samples are plotted to assess correlation in method for each patient. Trendlines indicate a linear regression fit with shaded 95% confidence intervals. Pearson’s correlations (R) are shown for all trendlines (*p < 0.05, ** p < 0.01, ***p < 0.001).

**Supplemental Figure 5.** Cell substate proportions for technical duplicate samples processed at single centers **(A)** and technical duplicate samples processed at both centers **(B)**. Samples from the same patient processed using different methods are shown next to each other; in (B), the corresponding pair of technical duplicates are shown sequentially. PRO denotes Cryo-PRO.

**Supplemental Figure 6.** Scatter plots of cell substate proportions from different processing sites. Each cell substate is represented by a different color. Proportion is the number of cells of one cell substate divided by the total number of cells of its parent cell type from that patient sample. The patient-paired Ficoll:Ficoll samples and Cryo-PRO:Cryo-PRO samples from the two different enrollment sites are plotted to assess correlation of technical duplicates for each patient. Trendlines indicate a linear regression fit with shaded 95% confidence intervals. Pearson’s correlations (R, *p < 0.05, ** p < 0.01, ***p < 0.001) are shown for all trendlines.

**Supplemental Figure 7.** Clonal expansion proportions for samples processed at single centers **(A)** and technical duplicate samples processed at both centers **(B)**. Samples from the same patient processed using different methods are shown next to each other. In (B), the corresponding pair of technical duplicates processed at the non-origin site are shown sequentially, the labeled site indicates where each sample was processed. PRO denotes Cryo-PRO.

**Supplemental Figure 8.** Scatter plot of the clonotype sequence frequency for all samples processed by different methods at single centers **(A)** and for technical duplicate samples processed at both centers by Ficoll **(B)** and Cryo-PRO **(C)**. The proportion of each unique sequence out of total TCR transcripts in each sample is plotted against its corresponding sample to compare similarities in Ficoll vs Cryo-PRO (A), or in processing center (B,C). Pearson’s correlations (R, *p < 0.05, ** p < 0.01, ***p < 0.001) are shown for all trendlines.

**Supplemental Figure 9.** Scatter plot of clonotype sequence frequency for all samples processed by different methods at single centers, individually plotted for each subject. The proportion of each unique sequence out of total TCR transcripts in each Ficoll sample is plotted against its corresponding Cryo-PRO sample to compare similarities in TCR clonal proportion.

**Supplemental Figure 10.** Scatter plot of the clonotype sequence frequency for technical duplicate samples processed at both centers by Ficoll (left) and Cryo-PRO (right), individually plotted for each subject. The proportion of each unique sequence out of total TCR transcripts in each MGH sample is plotted against its corresponding BIDMC sample to compare similarities in TCR clonal proportion.

**Supplemental Table 1.** Top marker genes by cell cluster and method. **(A)** Top 30 marker genes (ordered by highest average log_2_FC) for each cell type by method. **(B)** Top 30 marker genes (ordered by highest average log_2_FC) for each cell substate by method. Marker genes were excluded from the list if expressed in fewer than 25% of cells in that cluster (see Methods).

**Supplemental Table 2.** Differential gene expression by method. List of genes that were significantly (adjusted p value < 0.05) differentially expressed between methods for cell types and across all cells. Positive average log_2_FC indicates that the gene was enriched in Ficoll cells; negative average log_2_FC indicates that the gene was enriched in Cryo-PRO cells. Calculations were performed on pseudo-bulked samples (see Methods).

**Supplemental Table 3.** Fractional abundance of major cell types and transcriptional substates.

**Supplemental Table 4.** Phagocytosis assay results.

