## Supplemental figures for "Cryo-PRO facilitates whole blood cryopreservation for single-cell RNA sequencing of immune cells from clinical samples"

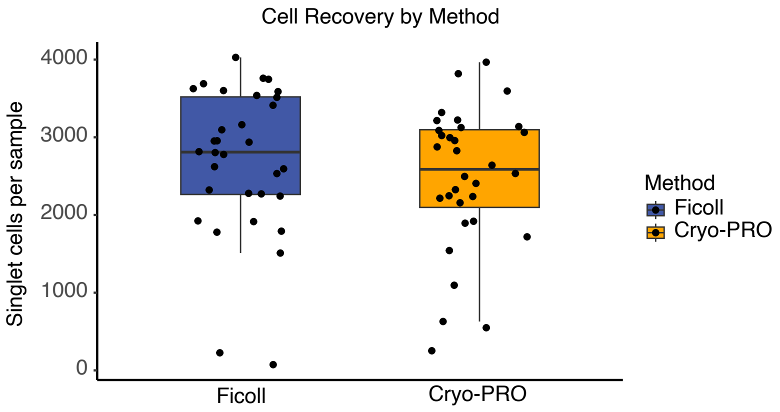


**Supplemental Figure 1.** Number of singlet cells sequenced per method. Starting blood sample volume was variable in Ficoll samples and was 1 mL in Cryo-PRO samples.

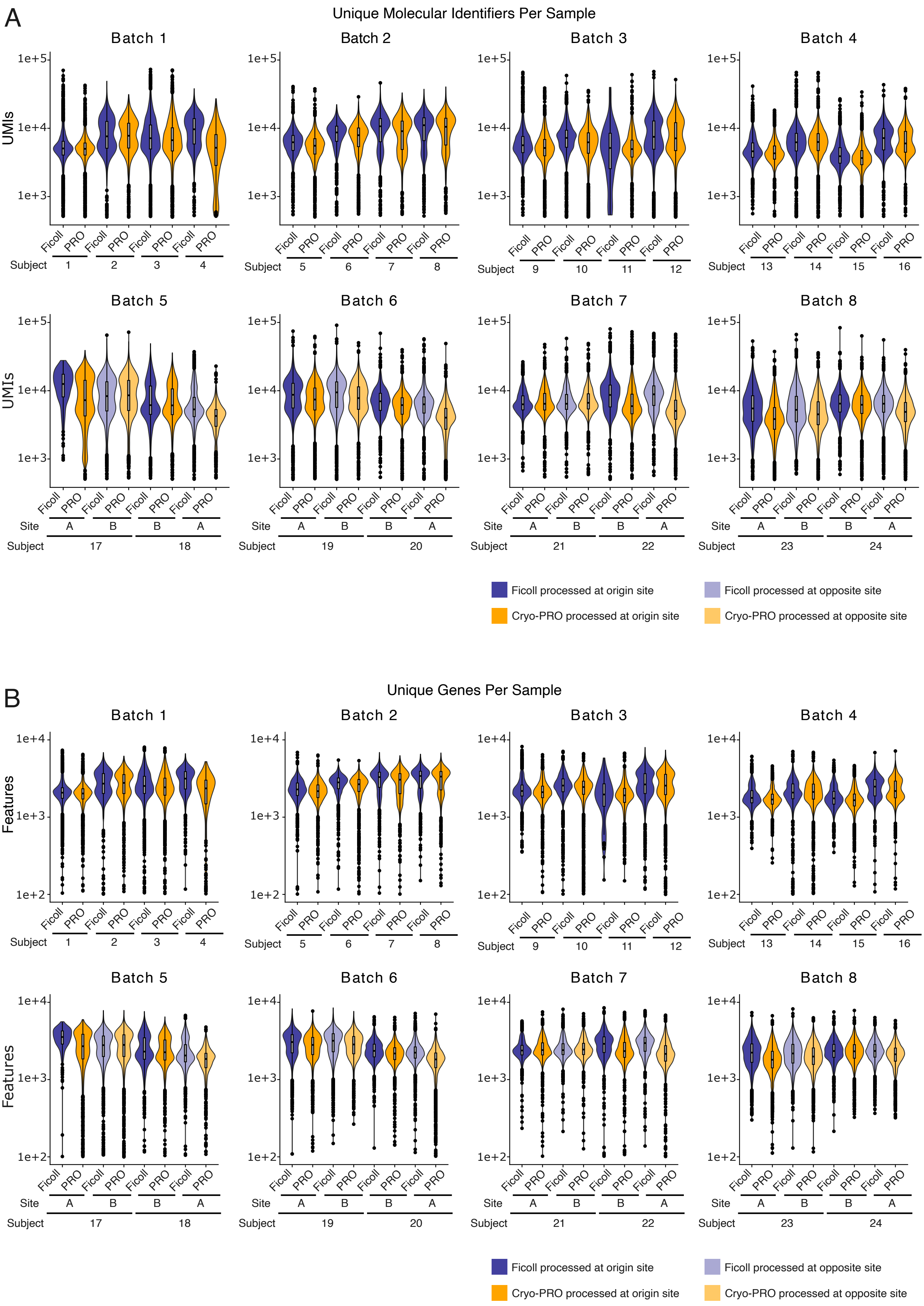
A
Unique Molecular Identifiers Per Sample
Batch 1
Batch 2
Batch 3
Batch 4
Batch 5
Batch 6
Batch 7
Batch 8
UMIs
1e+5
1e+4
1e+3
Ficoll
PRO
Subject
Site
1 2 3 4
5 6 7 8
9 10 11 12
13 14 15 16
17 18
19 20
21 22
23 24
A B B A
Ficoll processed at origin site
Ficoll processed at opposite site
Cryo-PRO processed at origin site
Cryo-PRO processed at opposite site
B
Unique Genes Per Sample
Features
1e+4
1e+3
1e+2

C

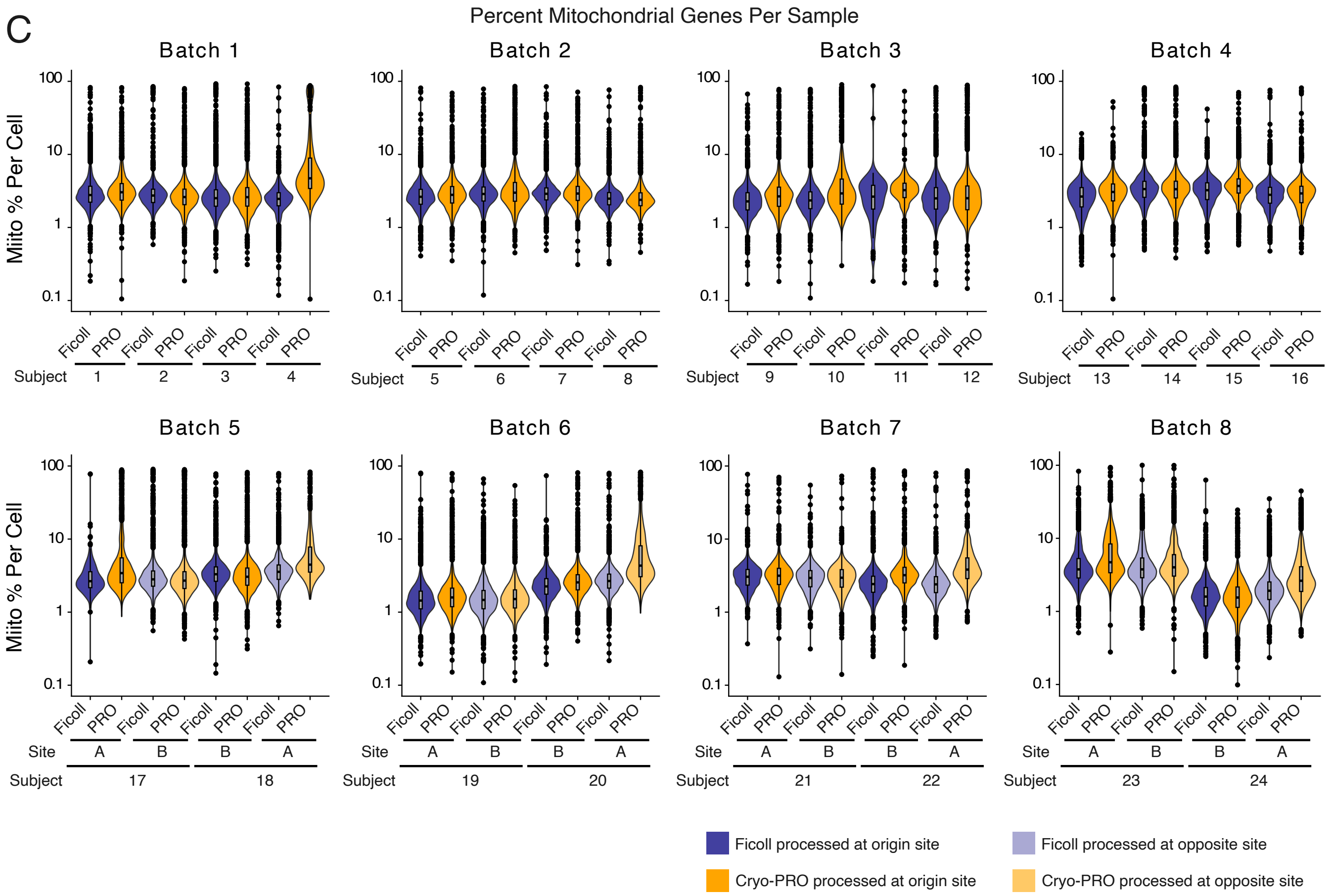
Percent Mitochondrial Genes Per Sample
Batch 1
Batch 2
Batch 3
Batch 4
Batch 5
Batch 6
Batch 7
Batch 8
Mito % Per Cell
100
10
1
0.1
Ficoll
PRO
Site
A
B
Subject
1 2 3 4 5 6 7 8 9 10 11 12 13 14 15 16
17 18 19 20 21 22 23 24
Ficoll processed at origin site
Ficoll processed at opposite site
Cryo-PRO processed at origin site
Cryo-PRO processed at opposite site

D

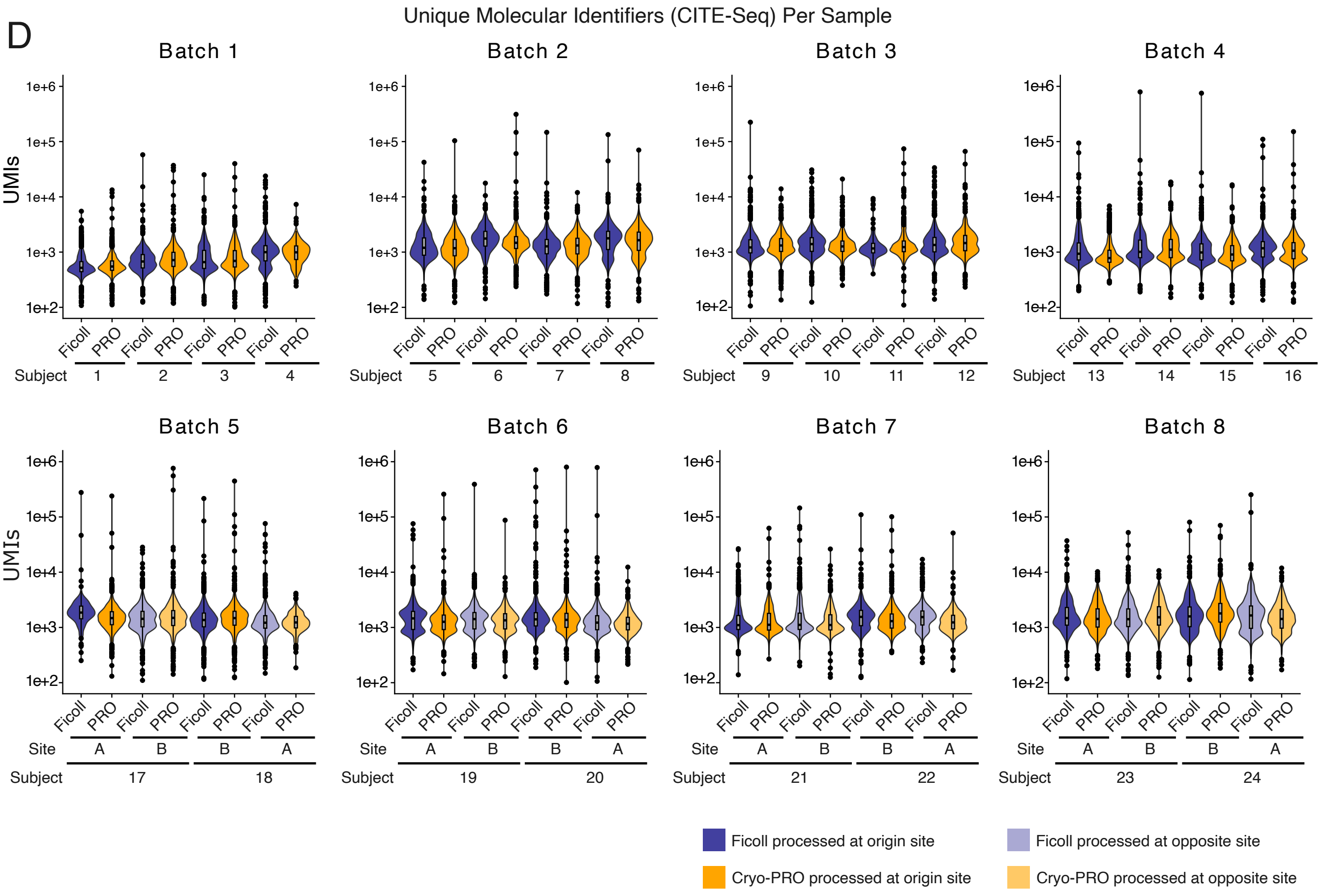
Unique Molecular Identifiers (CITE-Seq) Per Sample
Batch 1
Batch 2
Batch 3
Batch 4
Batch 5
Batch 6
Batch 7
Batch 8
UMIs
1e+6
1e+5
1e+4
1e+3
1e+2
Ficoll
PRO
Site
A
B
Subject
1 2 3 4 5 6 7 8 9 10 11 12 13 14 15 16
17 18 19 20 21 22 23 24
Ficoll processed at origin site
Ficoll processed at opposite site
Cryo-PRO processed at origin site
Cryo-PRO processed at opposite site

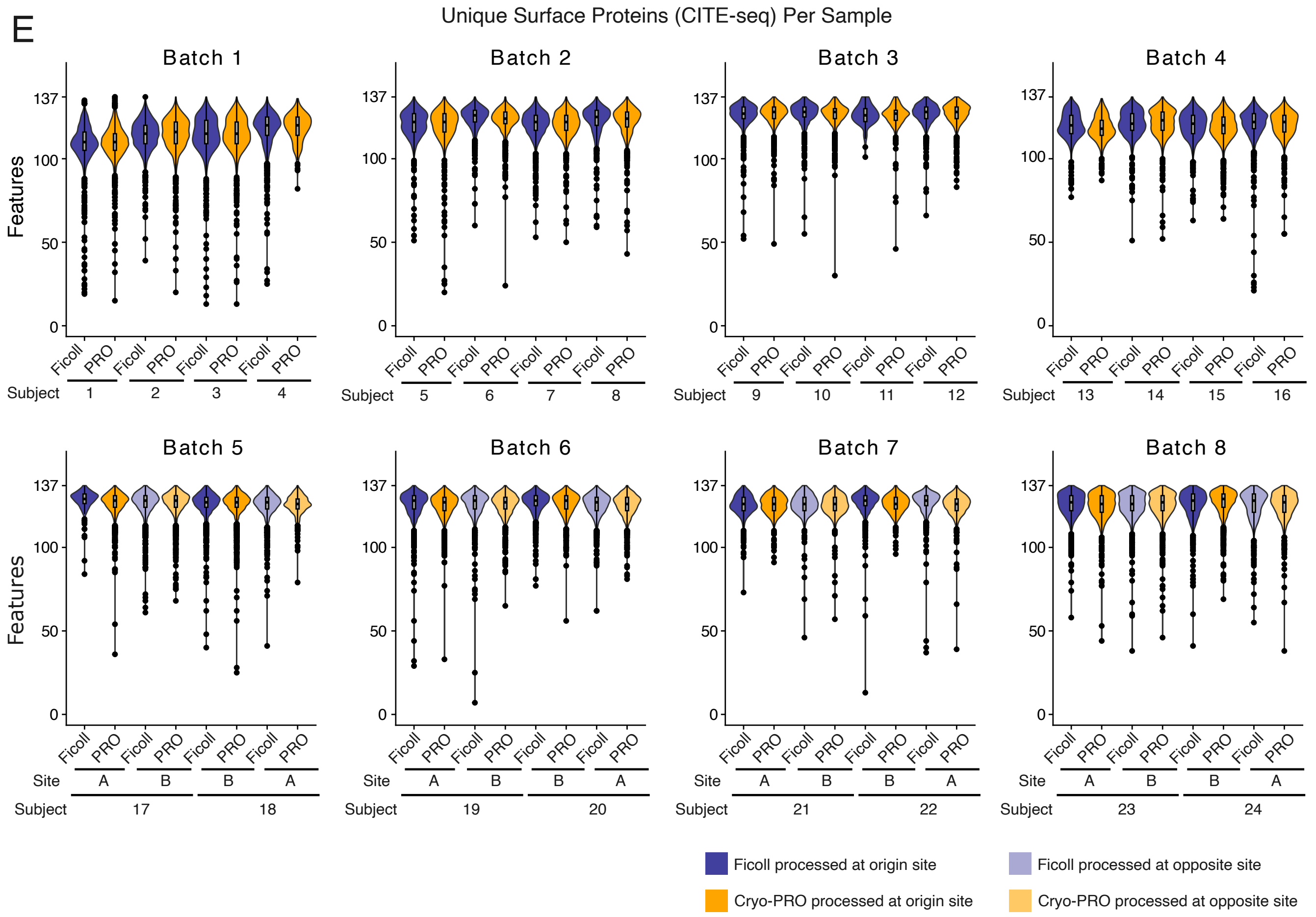


**Supplemental Figure 2.** Per-sample distributions of **(A)** UMIs of RNA transcripts, **(B)** unique genes, **(C)** percentage of mitochondrial transcripts, **(D)** unique surface protein features detected via CITE-seq, and **(E)** UMIs of surface protein features detected via CITE-seq per cell. Batches represent samples that were thawed, processed and sequenced together. Ficoll and Cryo-PRO samples from the same patient are plotted next to each other. For patients where parallel processing occurred at both clinical sites (bottom rows), the samples processed at the opposite site of enrollment are shown in lighter shades. A total of 137 different surface proteins were queried in the CITE-seq analysis. Violin plots show the distribution of each metric detected per cell; embedded boxplots indicate the median and IQR (whiskers = 1.5x IQR). PRO denotes Cryo-PRO.

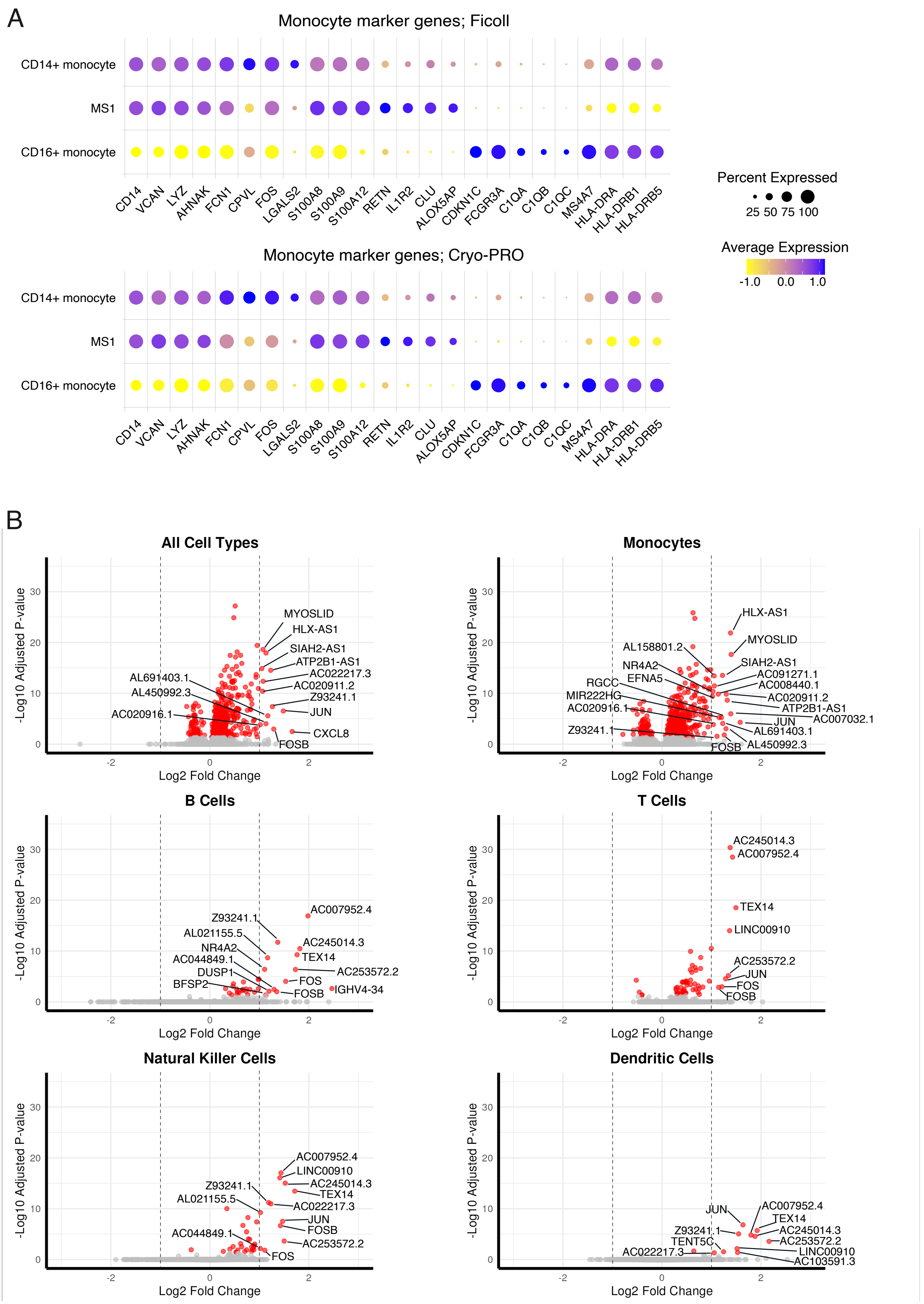
A
Monocyte marker genes; Ficoll
CD14+ monocyte
MS1
CD16+ monocyte
CD14 VCAN LYZ AHNAK FCN1 CPVL FOS LGALS2 S100A8 S100A9 S100A12 RETN IL1R2 CLU ALOX5AP CDKN1C FCGR3A C1QA C1QB C1QC MS4A7 HLA-DRA HLA-DRB1 HLA-DRB5
Monocyte marker genes; Cryo-PRO
CD14+ monocyte
MS1
CD16+ monocyte
CD14 VCAN LYZ AHNAK FCN1 CPVL FOS LGALS2 S100A8 S100A9 S100A12 RETN IL1R2 CLU ALOX5AP CDKN1C FCGR3A C1QA C1QB C1QC MS4A7 HLA-DRA HLA-DRB1 HLA-DRB5
Percent Expressed
25 50 75 100
Average Expression
-1.0 0.0 1.0
B
All Cell Types
MYOSLID
HLX-AS1
SIAH2-AS1
ATP2B1-AS1
AC022217.3
AC020911.2
Z93241.1
JUN
CXCL8
FOSB
AL691403.1
AL450992.3
AC020916.1
Monocytes
HLX-AS1
MYOSLID
SIAH2-AS1
AC091271.1
AC008440.1
AC020911.2
ATP2B1-AS1
AC007032.1
JUN
AL691403.1
AL450992.3
FOSB
AL158801.2
NR4A2
EFNA5
RGCC
MIR222HG
AC020916.1
Z93241.1
B Cells
AC007952.4
Z93241.1
AL021155.5
NR4A2
AC044849.1
DUSP1
BFSP2
AC245014.3
TEX14
AC253572.2
FOS
FOSB
IGHV4-34
T Cells
AC245014.3
AC007952.4
TEX14
LINC00910
AC253572.2
JUN
FOS
FOSB
Natural Killer Cells
AC007952.4
LINC00910
AC245014.3
TEX14
AC022217.3
Z93241.1
AL021155.5
JUN
FOSB
AC044849.1
AC253572.2
FOS
Dendritic Cells
JUN
AC007952.4
TEX14
AC245014.3
AC253572.2
LINC00910
AC103591.3
Z93241.1
TENT5C
AC022217.3
-Log10 Adjusted P-value
0 10 20 30
Log2 Fold Change
-2 0 2

**Supplemental Figure 3.** Dot plots of marker gene expression by each monocyte substate **(A)**. Color represents scaled relative expression (blue = higher expression). Size represents the proportion of cells in each substate where the feature was detected. Volcano plots showing genes differentially up-regulated (positive Log2FC) or down-regulated (negative Log2FC) in Ficoll compared to Cryo-PRO after pseudobulk analysis **(B)**. Genes with adjusted p-values of less than 0.05 are shown in red; those with $p < 0.05$ and abs(log2FC) > 1 are labeled. Plots are shown for differential gene expression among all cells (top left) and for each major cell type (subsequent plots).

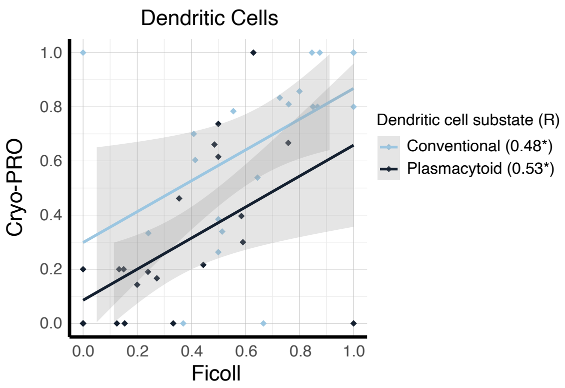


**Supplemental Figure 4.** Scatter plot of dendritic cell substate proportion from Ficoll and Cryo-PRO. Each point represents the proportion of one cell substate from one patient sample, as measured by each method. Each cell substate is represented by a different color. Proportion is the number of cells of one cell substate divided by the total number of dendritic cells from that patient sample. Patient-paired Ficoll:Cryo-PRO samples are plotted to assess correlation in method for each patient. Trendlines indicate a linear regression fit with shaded 95% confidence intervals. Pearson's correlations (R) are shown for all correlations (*$p < 0.05$, ** $p < 0.01$, ***$p < 0.001$).

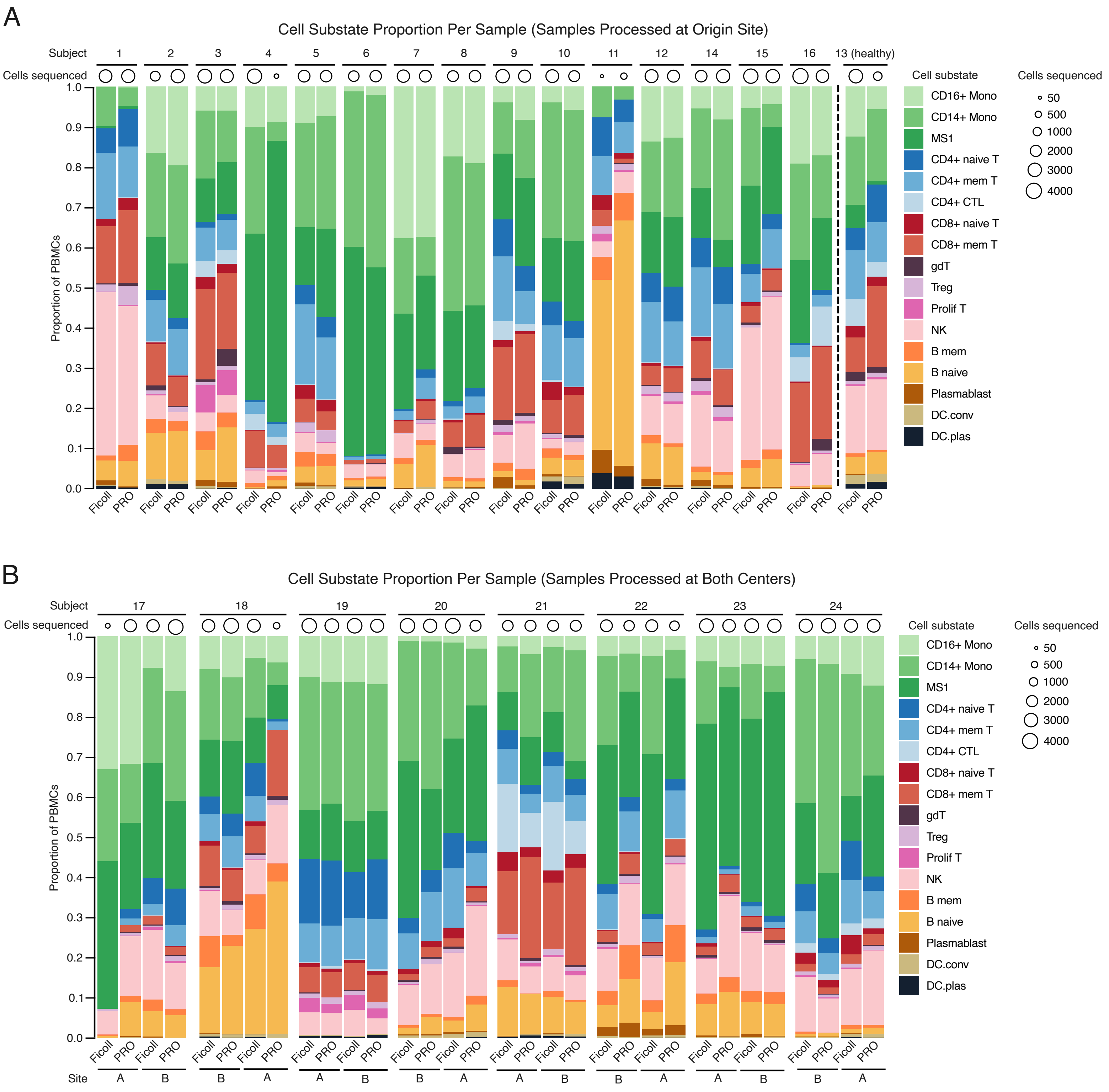


**Supplemental Figure 5.** Cell substate proportions for samples processed at single centers **(A)** and technical duplicate samples processed at both centers **(B)**. Samples from the same patient processed using different methods are shown next to each other. In (B), the corresponding pair of technical duplicates processed at the non-origin site are shown subsequently, with the labeled site indicating where each sample was processed. PRO denotes Cryo-PRO.

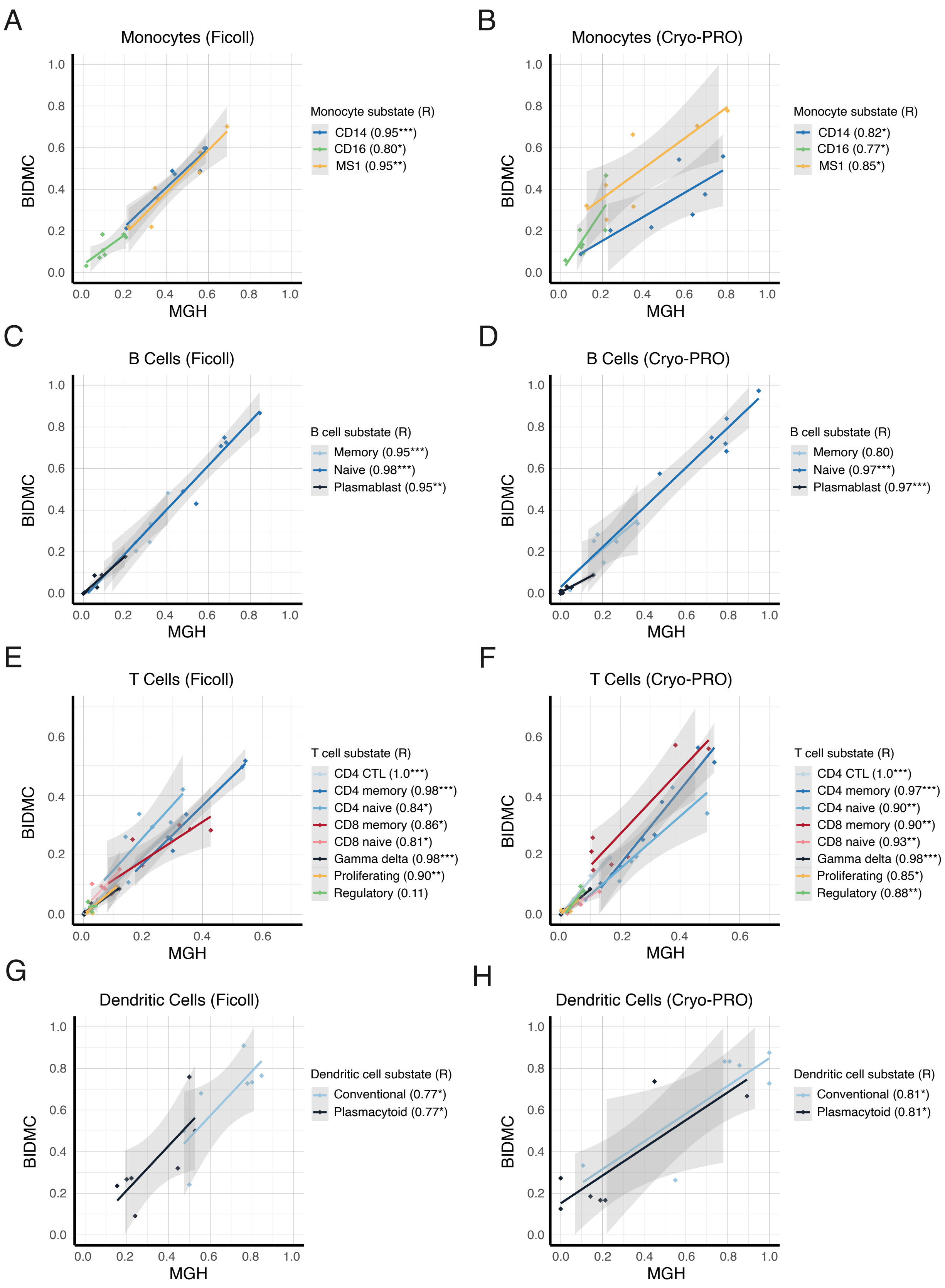


**Supplemental Figure 6**. Scatter plots of cell substate proportions from different processing sites. Each cell substate is represented by a different color. Proportion is the number of cellsof one cell substate divided by the total number of cells from its cell type from that patient sample. The patient-paired Ficoll:Ficoll samples and Cryo-PRO:Cryo-PRO samples from the two different enrollment sites are plotted to assess correlation of technical duplicates for each patient. Trendlines indicate a linear regression fit with shaded 95% confidence intervals. Pearson's correlations (R, *p < 0.05, ** p < 0.01, ***p < 0.001) are shown for all correlations.

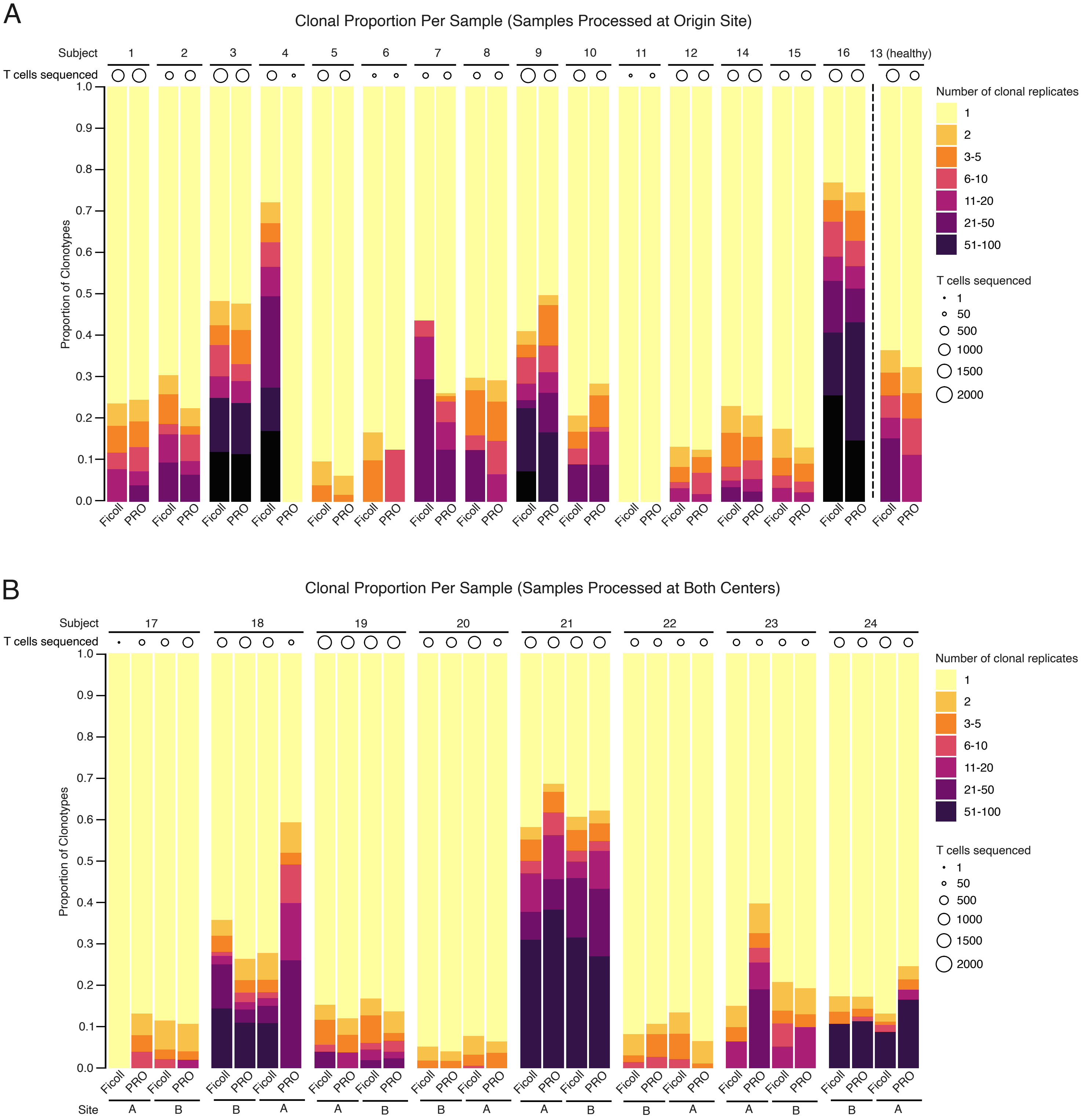


**Supplemental Figure 7.** Clonal expansion proportions for samples processed at single centers **(A)** and technical duplicate samples processed at both centers **(B)**. Samples from the same patient processed using different methods are shown next to each other. In (B), the corresponding pair of technical duplicates processed at the non-origin site are shown subsequently, with the labeled site indicating where each sample was processed. PRO denotes Cryo-PRO.

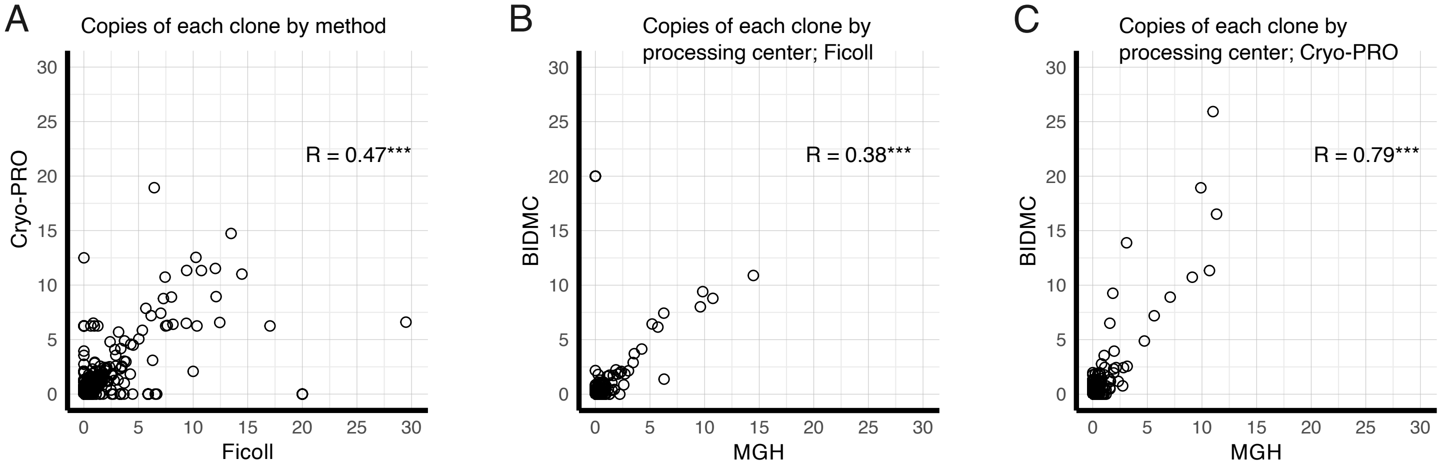


**Supplemental Figure 8**. Scatter plot of the clonotype sequence frequency for all samples processed by different methods at single centers **(A)** and for technical duplicate samples processed at both centers by Ficoll **(B)** and Cryo-PRO **(C)**. The proportion of each unique sequence out of total TCR transcripts in each sampleis plotted against its corresponding sample to compare similarities in Ficoll vs Cryo-PRO (A), or in processing center (B,C). Pearson's correlations (R, $*p < 0.05$, $** p < 0.01$, $***p < 0.001$) are shown for all correlations.

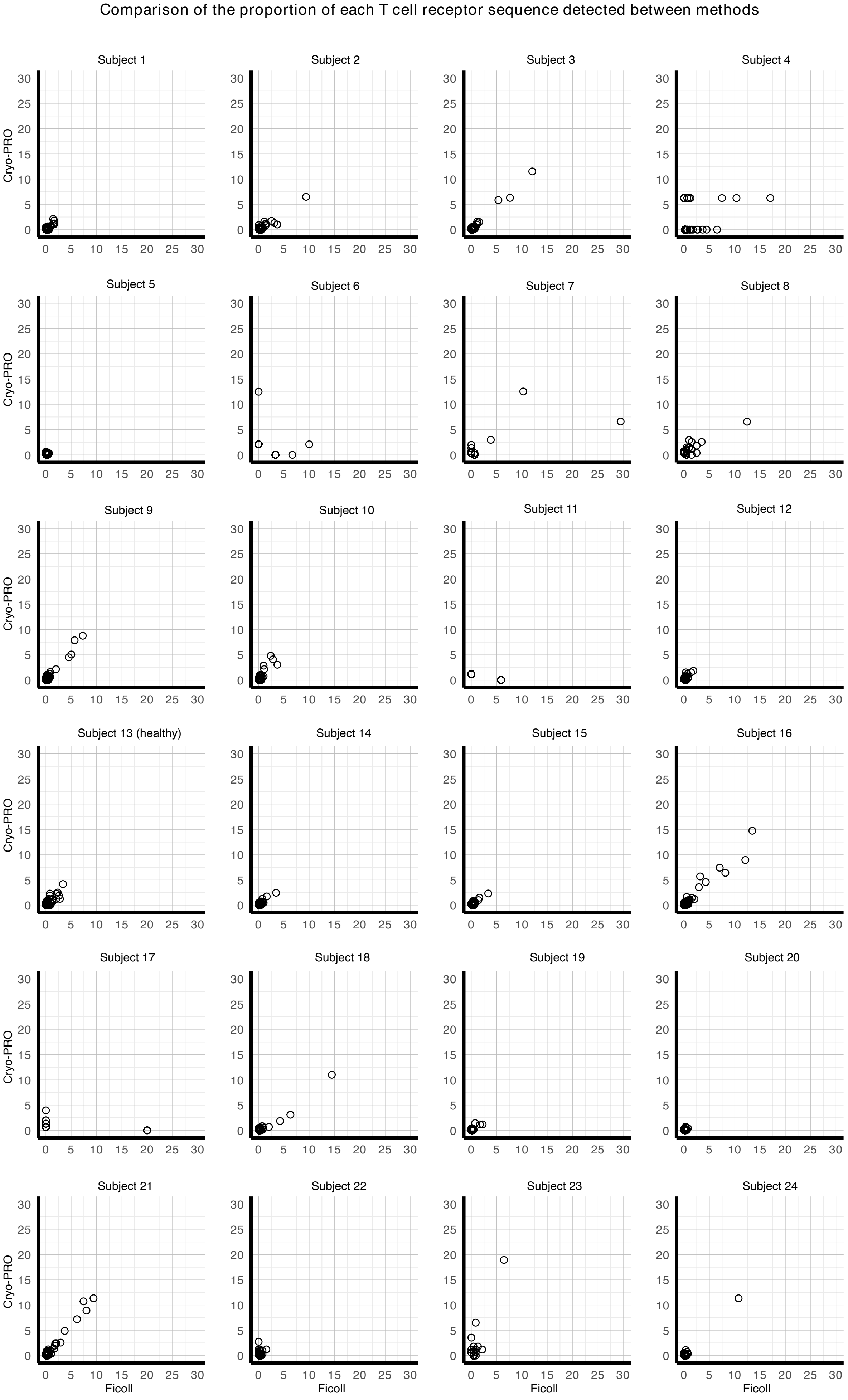


**Supplemental Figure 9.** Scatter plot of clonotype sequence frequency for all samples processed by different methods at single centers, individually plotted for each subject. The proportion of each unique sequence out of total TCR transcripts in each Ficoll sample is plotted against its corresponding Cryo-PRO sample to compare similarities in TCR clonal proportion.

Comparison of the proportion of each T cell receptor sequence detected between processing centers

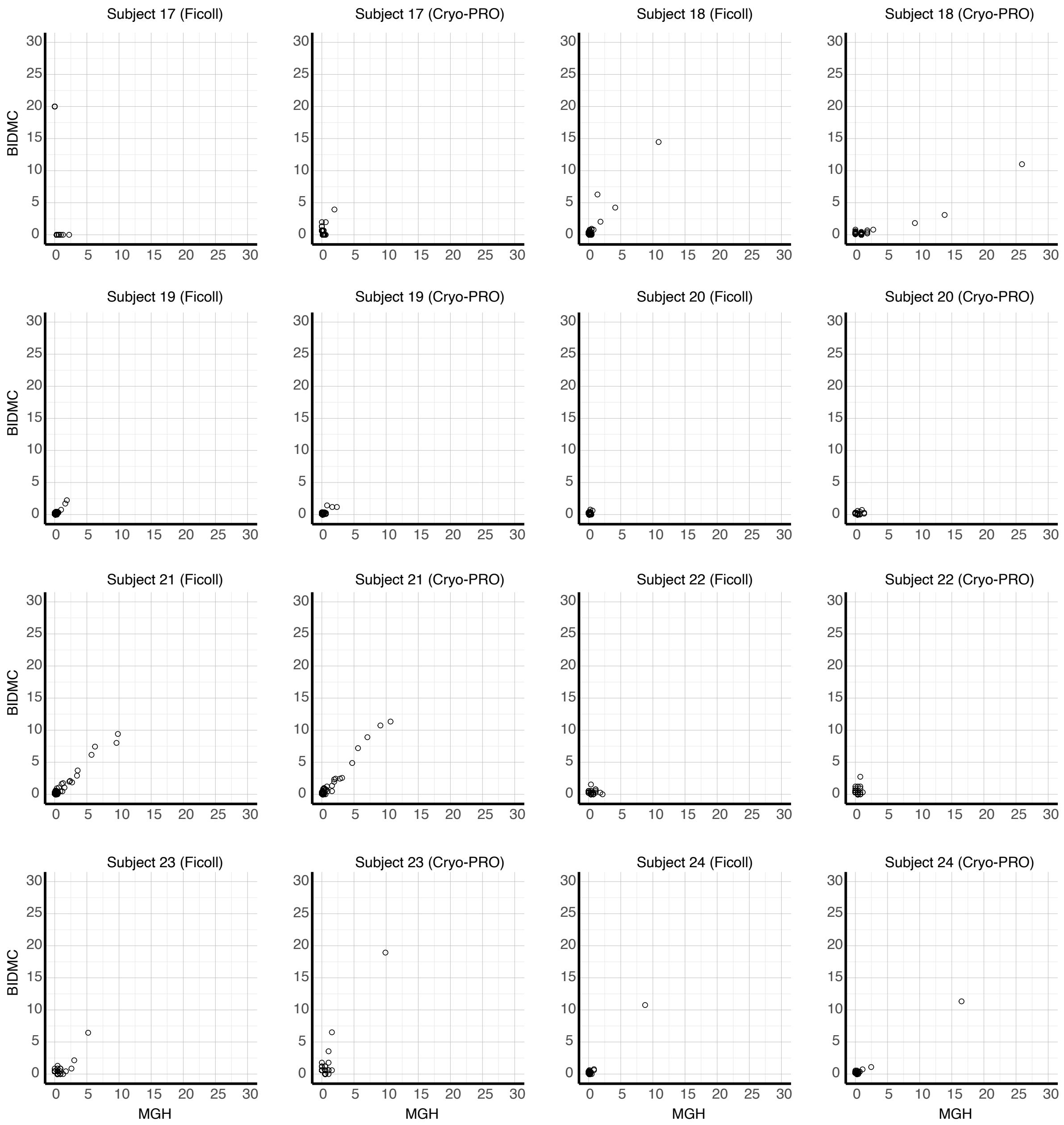


**Supplemental Figure 10.** Scatter plot of the clonotype sequence frequency for technical duplicate samples processed at both centers by Ficoll (left) and Cryo-PRO (right), individually plotted for each subject. The proportion of each unique sequence out of total TCR transcripts in each MGH sample is plotted against its corresponding BIDMC sample to compare similarities in TCR clonal proportion.
